# The Pulse of Artificial Intelligence in Cardiology: A Comprehensive Evaluation of State-of-the-Art Large Language Models for Potential Use in Clinical Cardiology

**DOI:** 10.1101/2023.08.08.23293689

**Authors:** Andrej Novak, Ivan Zeljković, Fran Rode, Ante Lisičić, Iskra A. Nola, Nikola Pavlović, Šime Manola

## Abstract

**Introduction:** Over the past two years, the use of Large Language Models (LLMs) in clinical medicine has expanded significantly, particularly in cardiology, where they are applied to ECG interpretation, data analysis, and risk prediction. This study evaluates the performance of five advanced LLMs—Google Bard, GPT-3.5 Turbo, GPT-4.0, GPT-4o, and GPT-o1-mini—in responding to cardiology-specific questions of varying complexity.

**Methods:** A comparative analysis was conducted using four test sets of increasing difficulty, encompassing a range of cardiovascular topics, from prevention strategies to acute management and diverse pathologies. The models’ responses were assessed for accuracy, understanding of medical terminology, clinical relevance, and adherence to guidelines by a panel of experienced cardiologists.

**Results:** All models demonstrated a foundational understanding of medical terminology but varied in clinical application and accuracy. GPT-4.0 exhibited superior performance, with accuracy rates of 92% (Set A), 88% (Set B), 80% (Set C), and 84% (Set D). GPT-4o and GPT-o1-mini closely followed, surpassing GPT-3.5 Turbo, which scored 83%, 64%, 67%, and 57%, and Google Bard, which achieved 79%, 60%, 50%, and 55%, respectively. Statistical analyses confirmed significant differences in performance across the models, particularly in the more complex test sets. While all models demonstrated potential for clinical application, their inability to reference ongoing clinical trials and some inconsistencies in guideline adherence highlight areas for improvement.

**Conclusion:** LLMs demonstrate considerable potential in interpreting and applying clinical guidelines to vignette-based cardiology queries, with GPT-4.0 leading in accuracy and guideline alignment. These tools offer promising avenues for augmenting clinical decision-making but should be used as complementary aids under professional supervision.

## Introduction

Over the last several years, we have witnessed a surge in the utilization of large language models (LLMs) for diverse applications, ranging from basic engineering problems to complex issues in medical research and its applications. LLMs are artificial neural networks typically built with transformer-based architecture that autonomously learn from data and can produce sophisticated and seemingly intelligent writing after being trained on a massive dataset (1). These models exemplify the adaptability of LLMs, requiring minimal reconfiguration to excel across numerous domains and tasks (2). Natural language processing powered by pre-trained language models is the key technology behind medical artificial intelligence (AI) systems that utilize clinical narratives (3).

There are numerous potential applications of LLMs in medicine, including support in clinical decision-making, knowledge retrieval, summarizing key diagnostic findings, triaging patients’ primary care concerns, enhancing patient health literacy, and more (2). They have great potential to modernize academic research by accelerating data analysis, literature reviews, and referencing (4–6). LLMs can assist with repetitive hospital tasks, such as writing discharge letters and analyzing large datasets to identify patterns, risk factors, and outcome predictions (3,7–9). In pursuit of deploying these advanced models into real-world clinical settings, we developed a prototype mobile application, ‘Dubravka,’ that integrates user-specific medical data and interfaces with a Large Language Model. This application is intended to serve as a stepping stone towards personalized, guideline-oriented patient care within the cardiology domain (10–18).

A notable achievement of GPT has been its performance on the United States Medical Licensing Examination (USMLE), where it attained scores at or near the passing threshold for all three components of the exam (19). This study assessed GPT’s clinical reasoning capabilities through its responses to the standardized questions of the USMLE, which emulate aspects of clinical decision-making. In subsequent research (20), GPT was evaluated using the American Heart Association (AHA) Basic Life Support (BLS) and Advanced Cardiovascular Life Support (ACLS) examinations. Although it initially fell short of the passing threshold, GPT’s responses were more pertinent and aligned with established guidelines compared to other AI systems, also offering reasoned explanations for its answers. Another study explored GPT’s proficiency in responding to queries from the Ophthalmic Knowledge Assessment Program (OKAP) exam (21). The model demonstrated significant accuracy in two parallel exams, although its performance varied across different ophthalmic subspecialties. It excelled in general ophthalmology while showing modest results in specialized areas such as neuro-ophthalmology and intraocular tumors. These findings, coupled with the heterogeneous results across various topics within the same specialty, have partly motivated the current study.

AI has been used in cardiology for some time, with computers interpreting electrocardiograms (ECGs) daily (22–24). A 2019 study showed that machine learning (ML) algorithms exceeded the diagnostic accuracy of a team of practicing cardiologists in classifying 12 heart rhythm types from single-lead ECGs, and ML can even be used to predict pathologies from ECG recordings (25, 26). However, the use of LLMs for ECG interpretation remains scarce. A study from early 2023 (27) evaluated the appropriateness of ChatGPT’s responses to questions regarding the prevention of cardiovascular diseases. The questions were crafted to mirror typical inquiries likely to be raised by individuals without a healthcare background. The majority (84%) of answers provided by ChatGPT were graded as appropriate. The study suggests the potential for LLMs to enhance education, improve patient health literacy, and provide counseling on the basics of cardiovascular prevention. Another recent study highlighted ChatGPT’s potential as an assisted decision support tool for straightforward clinical questions, though it was less effective for questions where general practitioners needed the help of cardiologists in decision-making (27,28).

Finally, let us mention a recent study (29) in which the performance of GPT-3.5 and GPT-4 was evaluated on the Polish Medical Final Examination (MFE) in both English and Polish. GPT-4 outperformed GPT-3.5, achieving mean accuracies of 79.7% in both languages and passing all versions of the MFE. In contrast, GPT-3.5 showed lower accuracies and was less consistent in passing the exams. Despite GPT-4’s superior performance, its scores were mostly lower than the average scores of medical students. A significant correlation was observed between the correctness of the answers and the index of difficulty for both models.

The primary aim of this study was to evaluate the clinical utility of five State-of-the-Art Large Language Models (LLMs)—Google Bard, GPT-3.5 Turbo, GPT-4.0, GPT-4o, and GPT-o1-mini—by comparing their performance on a series of cardiology-focused clinical vignettes. These vignettes were designed to represent a spectrum of scenarios, from basic cardiovascular care to clinical decision-making. Our objective was to assess each model’s accuracy, understanding of medical terminology, ability to provide clinically relevant and contextually appropriate responses, and adherence to guidelines. The overarching goal was to provide insights into the readiness of LLMs for application in real-world medical practice. To the best of our knowledge, the applications of LLMs in this context - spanning both vertical (across various levels of difficulty) and horizontal (comparing different LLMs on identical question sets) dimensions - have not been previously explored.

## Methods

In this cross-sectional study, we aimed to conduct a comparative analysis of five state-of-the-art LLMs: Google Bard (based on PaLM 2), GPT-3.5 Turbo, GPT-4.0 (model 0613), GPT-4o-mini (model gpt-4o-mini-2024-07-18), and GPT-4o (model gpt-4o-2024-08-06), all available as text-based chat interfaces provided by OpenAI and Google. These models were assessed based on four sets of vignette-based clinical scenarios, differentiated by their levels of complexity, and denoted as test sets A, B, C, and D, respectively. A significant portion of the questions featured five potential responses, among which only one was correct. However, a minor fraction of the queries deviated from this format, presenting three, four, six, or seven potential answers, yet still with only one correct answer.

The test set A is composed of 24 queries from the ANA exam (American Nurses Association), encompassing various clinical situations pertinent to fundamental cardiovascular patient care. Test set B includes 25 queries from the ACO exam (American College of Osteopathic Internists), encompassing an assortment of clinical circumstances and management strategies in cardiology centering around emergency states and coronary artery disease, along with scoring systems such as the Syntax Score. The test set C is a custom dataset formulated by the study authors, containing 60 questions that mirror the format and objectives of the USMLE Step 2 and 3 exams. Specifically, the set includes questions on general cardiology (18 questions), ischemic heart disease (12 questions), heart failure (9 questions), arrhythmia (12 questions), and a combination of questions dealing with valvular, endocardial, myocardial, and pericardial diseases (9 questions). Lastly, test set D incorporates 90 vignettes, including 15 questions from reference (30) and others created to follow the structure and outcomes of references (30, 31) while being aligned with current AHA guidelines. The questions featured different background scenarios, altered demographics, and reported anamnestic data, maintaining the core concepts of the board-type questions. Questions in both Test set C and Test set D were reviewed and cross-validated by a panel of four senior cardiologists to ensure clinical accuracy, relevance, and appropriate complexity.

Test sets B, C, and D feature various patient demographics. Questions exclusively pertaining to image data were excluded, and ECG or cardiac imaging graphical data supporting the vignette were incorporated into the questions textually, describing these findings following a standard format and terminology. Details of each question set, and the corresponding results are provided in Table 1.

**Table 1:** Comparative performance of Google Bard, GPT 3.5 Turbo, and GPT 4.0, GPT o1-mini and GPT-4o on four sets of questions with varied complexity in cardiology.

| Test | Level of difficulty | Google Bard | GPT-3.5 Turbo | GPT-4.0 | GPT-o1-mini | GPT-4o |
| --- | --- | --- | --- | --- | --- | --- |
| A – ANA | Basic (Cardiac nurse) | 19/24 | 20/24 | 22/24 | 21/24 | 21/24 |
| B - ACO | Mixed | 15/25 | 16/25 | 22/25 | 18/25 | 19/25 |
| C - Custom | USMLE-like Step 2/3 Question | 30/60 | 40/60 | 48/60 | 47/60 | 46/60 |
| D - Difficult | ABIM board reviews | 50/90 | 51/90 | 76/90 | 50/90 | 71/90 |

Note: Each entry denotes the number of correctly answered questions out of the total questions in each respective set. ANA - American Nurses Association; ACO - American College of Osteopathic Internists, USMLE - United States Medical Licensing Examination, ABIM - American Board of Internal Medicine cardiology board.

We utilized the text-based chat interfaces provided by OpenAI and Google for our inquiries and data collection. For the mobile application ‘Dubravka,’ which was developed as part of this research at Dubrava University Hospital, we employed the OpenAI GPT-3.5 Turbo API to generate patient-tailored cardiovascular prevention advice. The application, programmed in Flutter, stores basic patient information locally and provides them as hidden parameters to the LLM prompt, enabling personalization of outputs. The analysis was performed with the default temperature parameters set to 1 (on a scale with a minimum of 0 and a maximum of 2 in the OpenAI web application Playground) and 0.7 for Google Bard. The *top p* parameter was always set to 1 (default), as altering both parameters is not recommended^1^. The temperature parameter influences the randomness of the generated text (29). When set to a higher value, LLMs produce more diverse outputs by assigning equal probabilities to a broader range of words or tokens, leading to less predictable and more exploratory outputs. Conversely, a lower temperature setting makes the model more deterministic, favoring higher probability tokens and yielding more coherent, conservative outputs. Variations in the temperature parameter suggest that, within this range (min-to-max), the value primarily affects the overall diversity of the response output rather than the representation of medical knowledge encoded in the model (29). Prompts sent through the models consisted of the exact questions from the test sets, without additional comments or context. Each model’s final response was obtained and saved. Final answers from highlighted prompts were stored in Appendix 1.

The accuracy of all five models—GPT-4.0, GPT-3.5 Turbo, Google Bard, GPT-4o, and GPT-o1-mini—was calculated for each test by dividing the number of correct answers by the total number of questions in the respective test set. Performance differences among the models were analyzed using a Friedman test, a non-parametric alternative to repeated measures ANOVA, to account for the ranking nature of the data. Post-hoc pairwise comparisons were performed with Bonferroni correction to identify statistically significant differences between the models when the omnibus test indicated significance. The significance level for all tests was set at 0.05. Mobile application ‘Dubravka’ was programmed in Flutter, an open-source user interface software development kit. Additionally, Python 3, a high-level, general-purpose programming language, was employed for performing calculations and statistical inference through the computing platform Jupyter Notebook 6.4.12^2^.

The outputs generated from the LLMs were evaluated based on correct and incorrect responses. Incomplete answers or rare instances where the model yielded two answers as correct were treated as incorrect. The previously mentioned panel of four senior cardiologists (three interventional cardiologists and one imaging subspecialist) with over ten years of professional experience in the field, evaluated the answers, i.e., the full explanations given by the LLMs, following these criteria: accuracy, understanding of medical terminology, and clinical relevance. They also considered the depth of the perceived understanding and explanation provided by the LLMs for each response. These were not scored numerically. The panel discussed their observations in detail.

The primary goal of this study was to appraise the clinical relevance of the responses generated by the LLMs in response to inquiries seeking medical advice across the clinical continuum, including cardiovascular prevention, general cardiology, heart failure, arrhythmia, acute cardiovascular care management, and finally valvular, myocardial, endocardial and pericardial pathologies with a strong emphasis towards decision-making.

## Results

Our findings demonstrate that GPT-4.0 consistently delivered superior performance across all four sets of questions, achieving the highest scores in integrating clinical scenarios, adhering to current cardiology guidelines, and computing and interpreting various scores and indices. The GPT-4o and GPT-o1-mini models also performed well, following GPT-4.0 by a narrow margin, particularly excelling in USMLE-like Step 2/3 questions and ABIM board reviews. These models demonstrated strong reasoning abilities and reliable adherence to clinical guidelines, making them effective tools for complex medical queries. GPT-3.5 Turbo, while performing reasonably well, lagged behind the GPT-4 family of models, often resorting to speculative or less evidence-based answers, particularly in more difficult scenarios. On the other hand, Google Bard underperformed significantly across all test levels, frequently failing to provide direct answers or offering overly simplistic explanations that lacked depth and precision.

Despite their varying levels of accuracy, all models demonstrated an ability to provide responses that were perceived as correct and supplemented by explanatory commentary, often discounting alternative answers. Detailed performance data for each model across the different sets of questions are presented in Figure 1 and Table 1.

**Figure 1.**
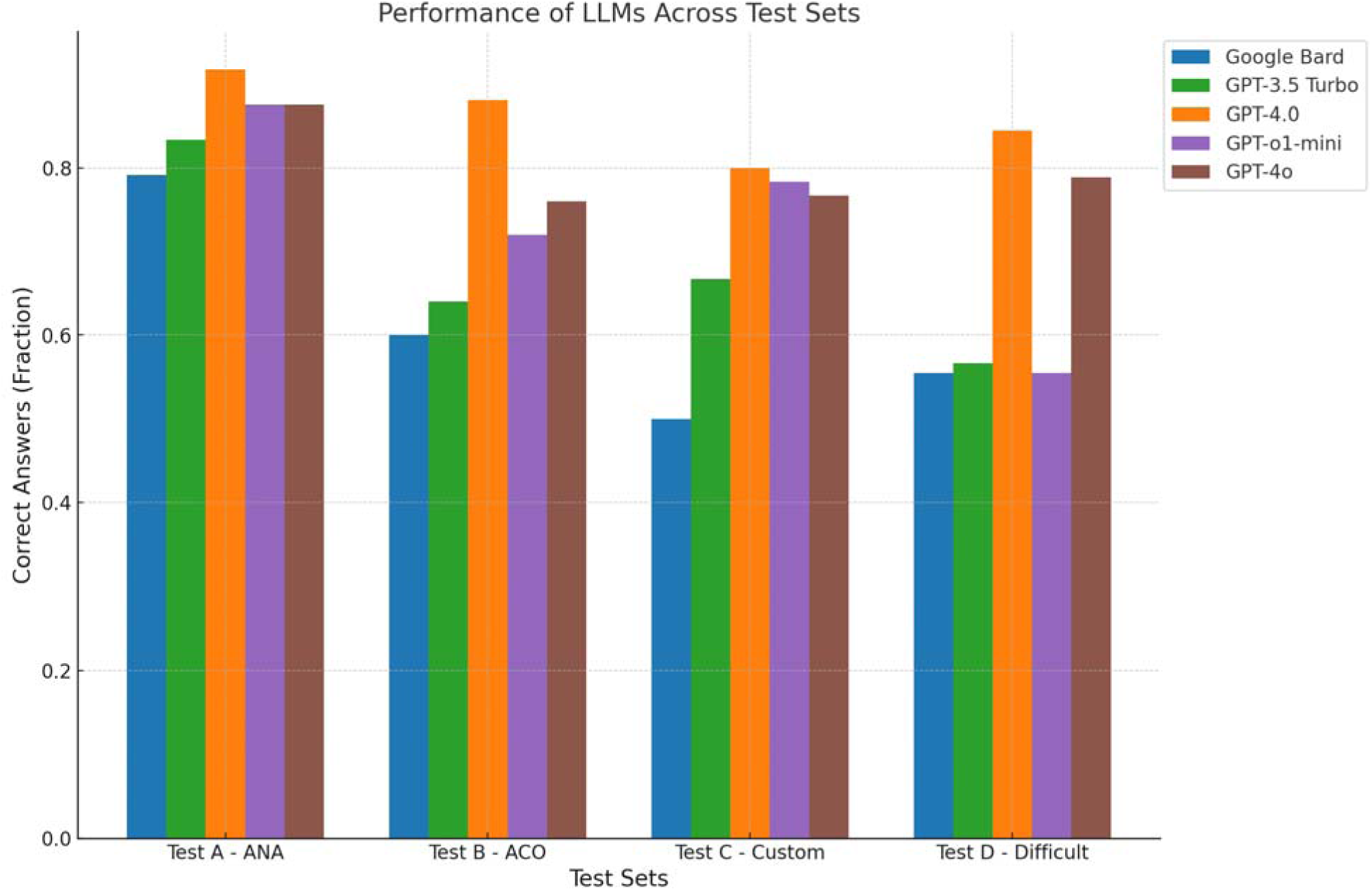
Model Performance Across Different Test Sets. The provided bar graph visually compares the performance of three different Large Language Models: Google Bard, GPT-3.5 Turbo, and GPT-4.0 across four distinct test sets, labeled A, B, C, and D. Each test set represents a different level of difficulty ranging from basic (Cardiac nurse, Test A) to difficult (Board reviews, Test D). The vertical axis of the graph denotes the accuracy percentage, quantifying the proportion of correctly answered questions. For each test set, the graph depicts three bars, each corresponding to one of the three LLMs. ANA - American Nurses Association exam queries; ACO - American College of Osteopathic Internists exam queries; Custom - custom dataset queries formulated by the study authors, that mirror the format and objectives of the United States Medical Licensing Examination (USMLE) Step 2 and 3 exams; Difficult - incorporates 90 board-like queries created to follow the structure and outcomes of established American Board of Internal Medicine (ABIM) cardiology board review reference books (30, 31) while being aligned with current AHA guidelines.

### Accuracy

The performance statistics for each model are as follows: Test Set A (Basic Level): GPT-4.0 achieved a 92% success rate, with GPT-4o and GPT-o1-mini scoring 88% each. GPT-3.5 Turbo followed with 83%, while Google Bard scored 79%. Test Set B (Mixed Level): GPT-4.0 had an 88% success rate, higher than the 76% and 72% rates of GPT-4o and GPT-o1-mini, respectively. GPT-3.5 Turbo and Google Bard scored 64% and 60%, respectively. Test Set C (USMLE-like): GPT-4.0 achieved an 80% success rate, compared to 78% and 77% for GPT-4o and GPT-o1-mini. GPT-3.5 Turbo scored 67%, and Google Bard with 50%. Test Set D (Difficult Level): GPT-4.0 outperformed others with an 84% success rate, followed by GPT-4o at 79% and GPT-o1-mini at 76%. GPT-3.5 Turbo and Google Bard scored 57% and 55%, respectively.

### Understanding of Medical Terminology

Terms related to cardiology, including those pertaining to human anatomy, medical diagnoses, generic drug names, diagnostic tests, and laboratory parameters, were correctly identified by all models. GPT-4.0, GPT-4o, and GPT-o1-mini were the most accurate and proficient, consistently employing medical terminologies appropriately and aligning their answers with current clinical guidelines. GPT-3.5 Turbo demonstrated a solid understanding of the terminology and provided detailed explanations of each treatment option. However, its application of these terms within the framework of current clinical guidelines was occasionally flawed. Google Bard, by contrast, showed significant gaps in understanding medical terms, leading to multiple inaccurate statements and recommendations, as evidenced in Table 2 and Table 4.

**Table 2.**
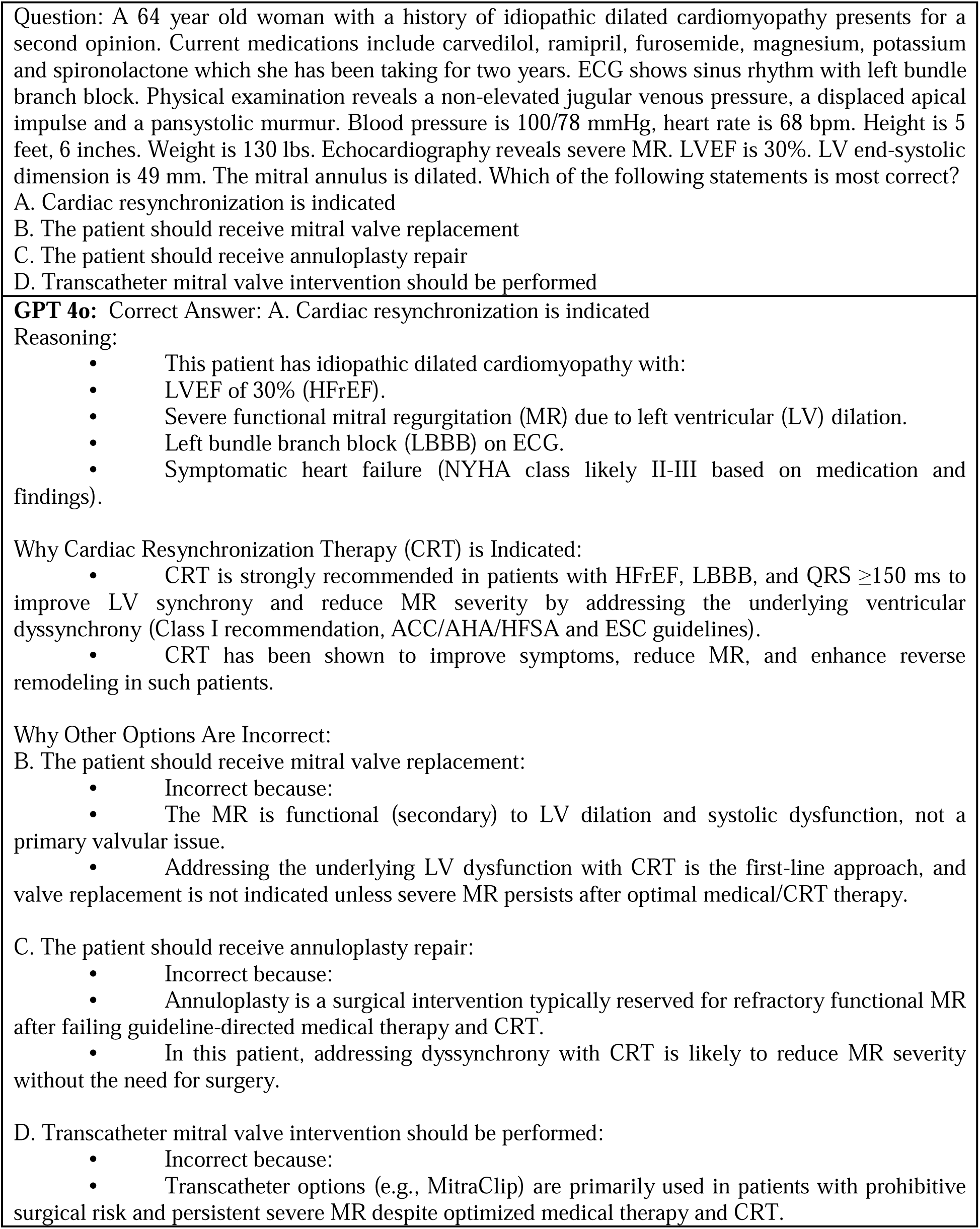
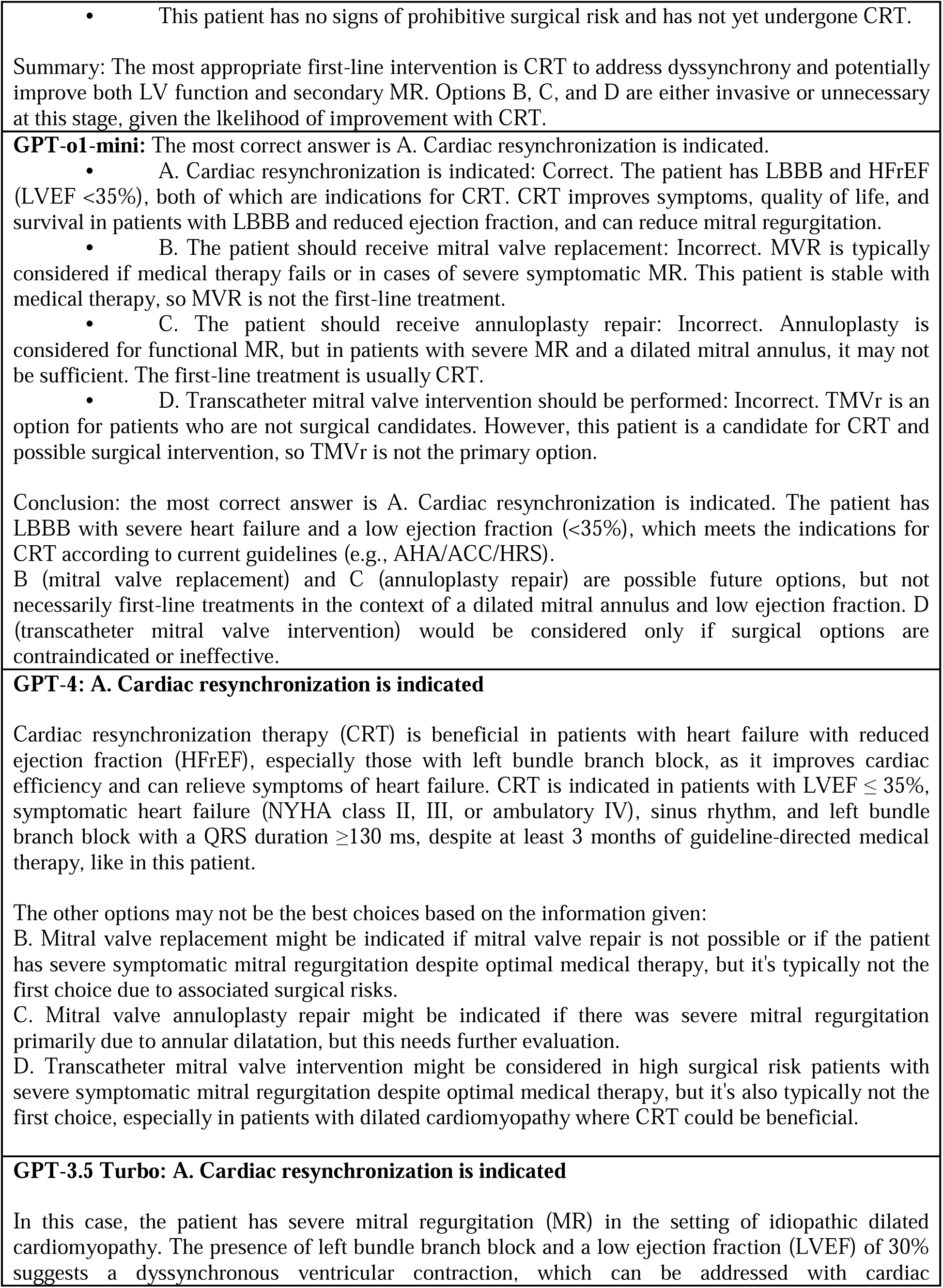
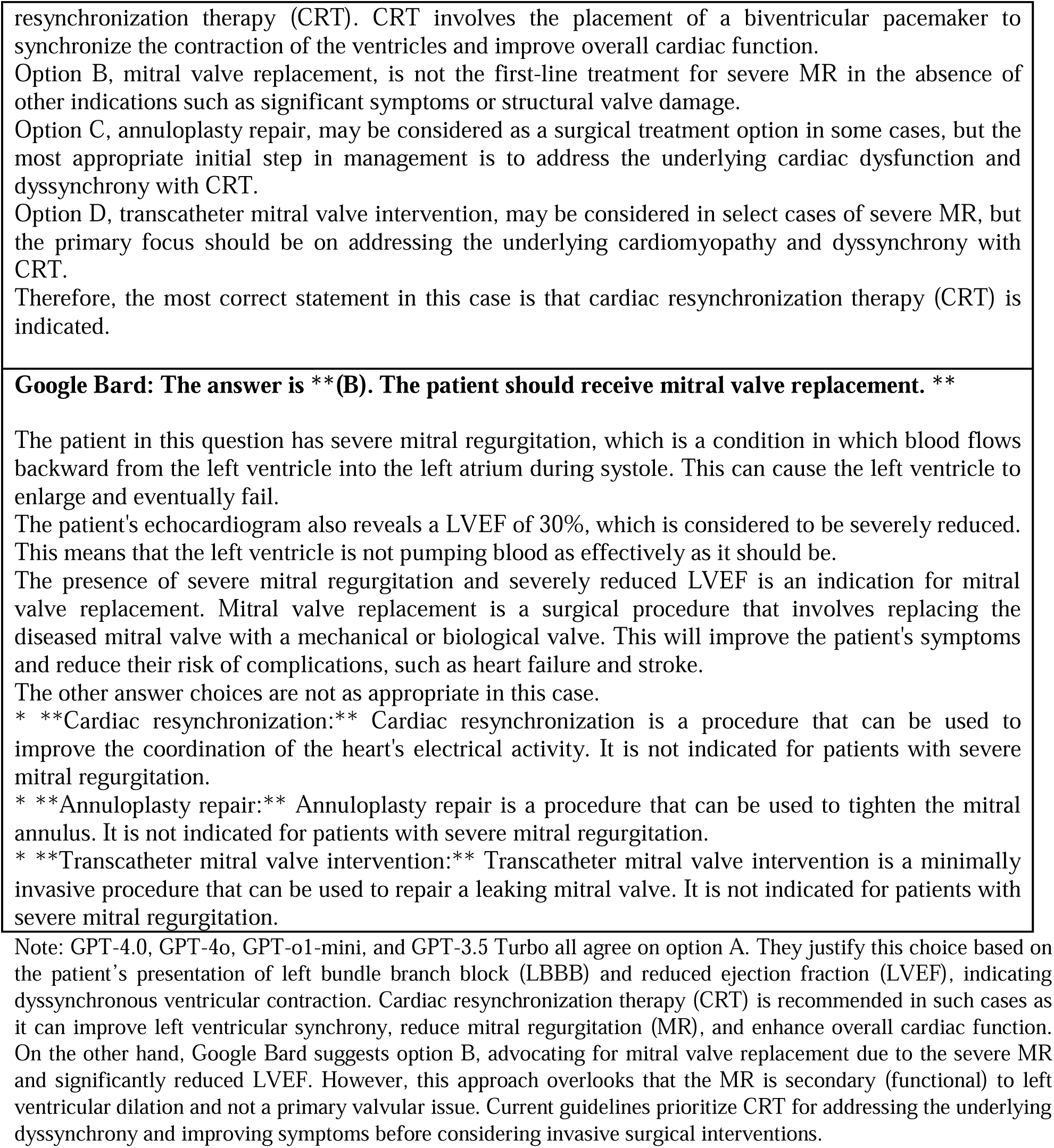
(Test set D, Question 3, Chapter 21 in. (**30**)) This table presents a case of a 64-year-old woman with idiopathic dilated cardiomyopathy, her clinical details, and a multiple-choice question on the most suitable treatment option.

### Clinical Relevance and Contextual Understanding

GPT-4.0, GPT-4o, and GPT-o1-mini demonstrated a strong understanding of clinical context and consistently adhered to established guidelines, including the 2018 AHA/ACC Guideline on the Management of Blood Cholesterol. These models occasionally referenced other notable guidelines, such as the ACC/AHA/HFSA guidelines for heart failure management and ESC guidelines for cardiac resynchronization therapy, aligning their responses with evidence-based practices. However, none of the models cited specific expert papers or primary literature directly. GPT-3.5 Turbo, while generally detailed in its explanations, occasionally deviated from guideline recommendations, leading to less accurate clinical advice. Google Bard showed significant gaps in guideline adherence, providing inconsistent recommendations that often conflicted with established clinical standards (see Table 3, Table 5, and Table 6).

**Table 3.**
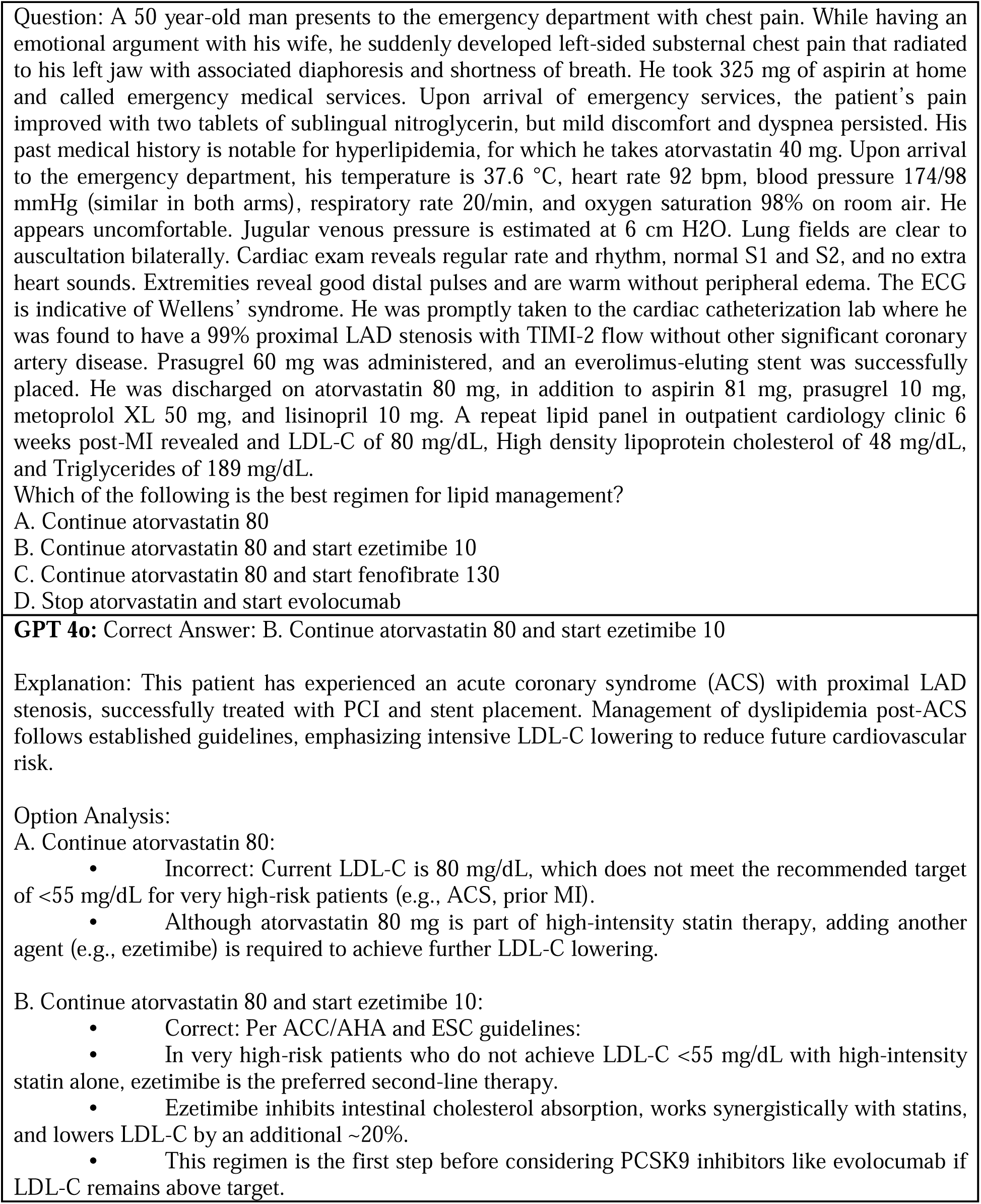
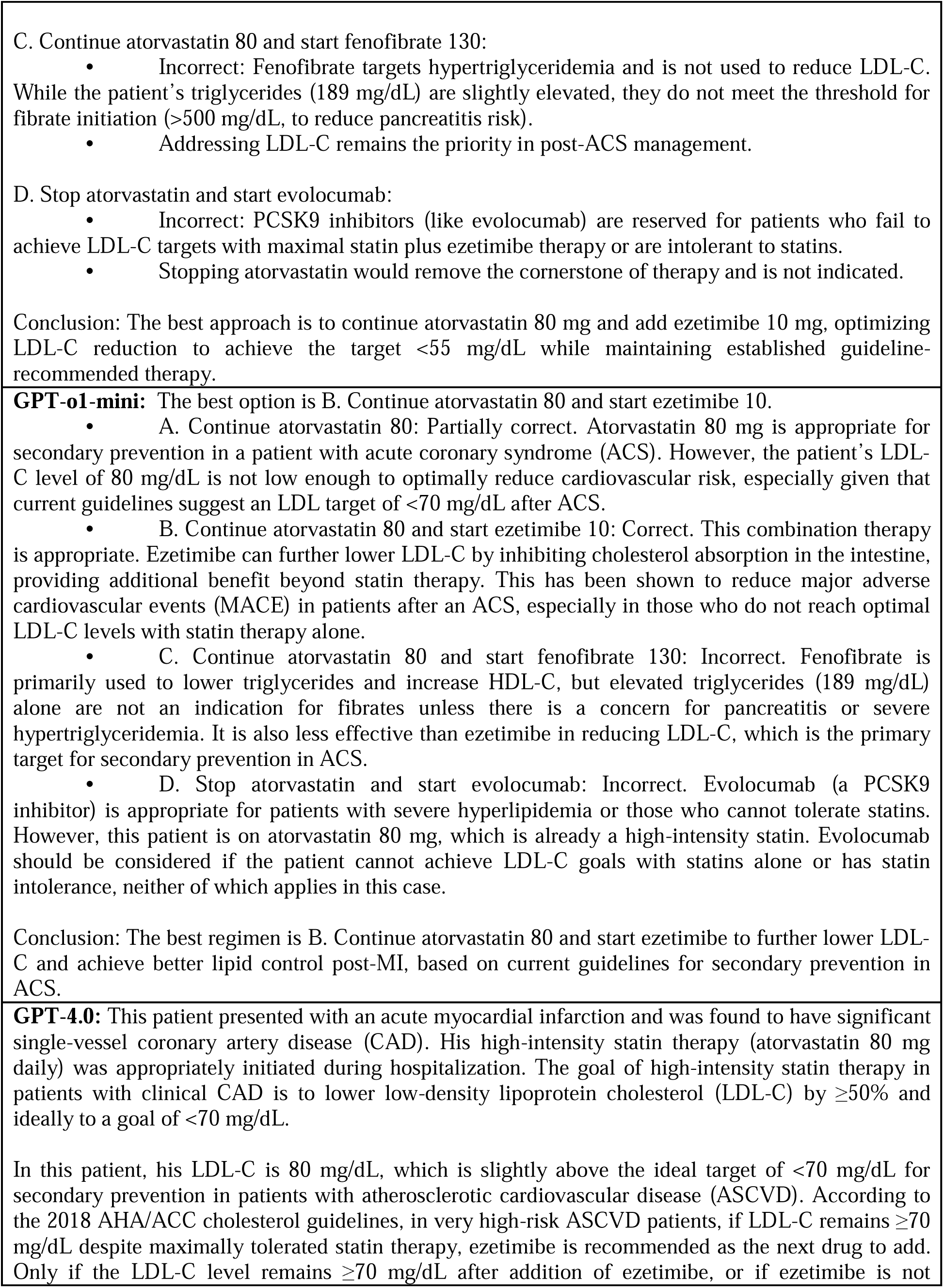
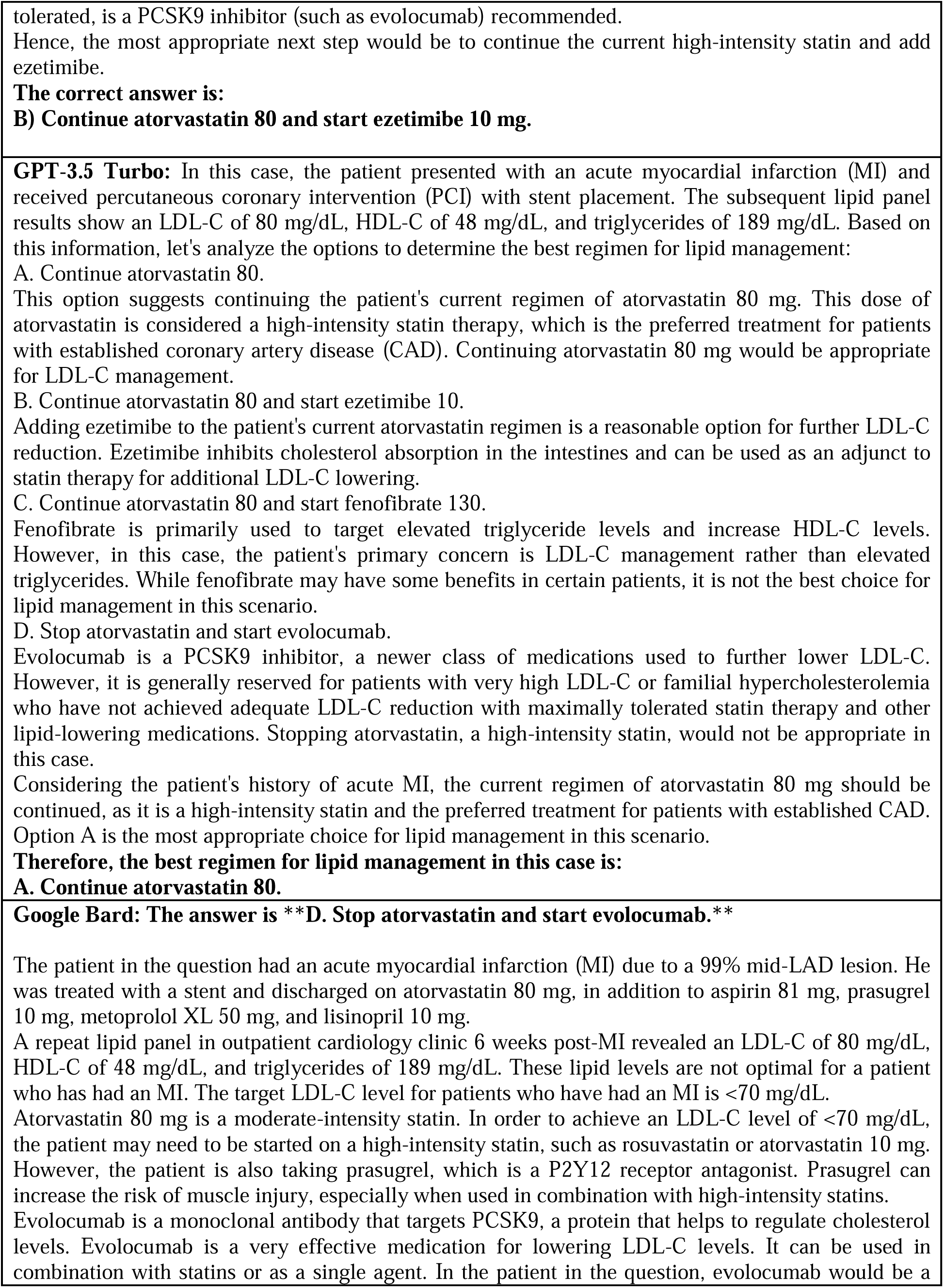
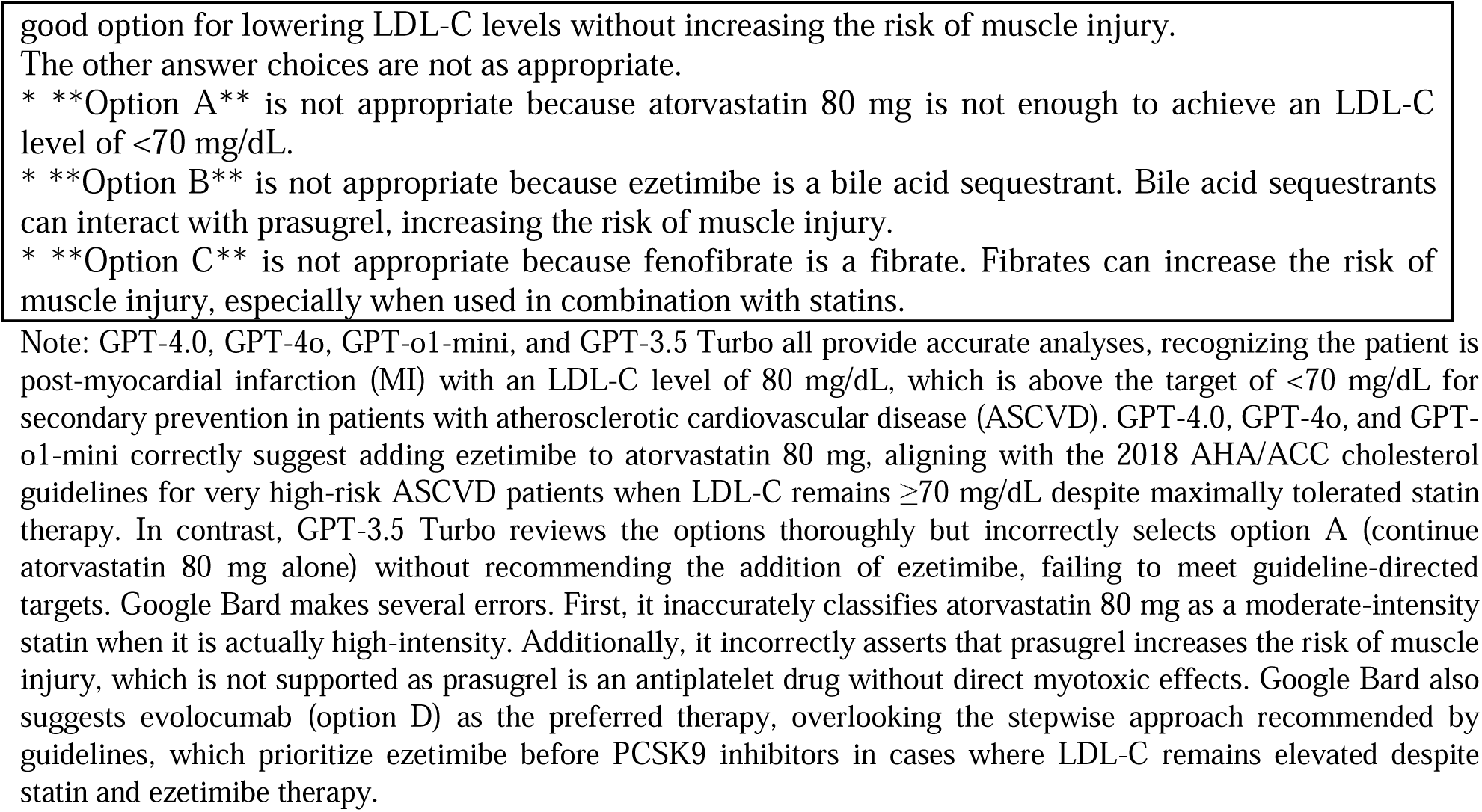
(Test set D, Question 4, Chapter 4 in. (**30**)) The table presents the clinical scenario of a 50-year-old man experiencing chest pain, detailing his medical history, symptoms, treatment, and follow-up. It concludes with a multiple-choice question regarding optimal lipid management strategy.

**Table 4.**
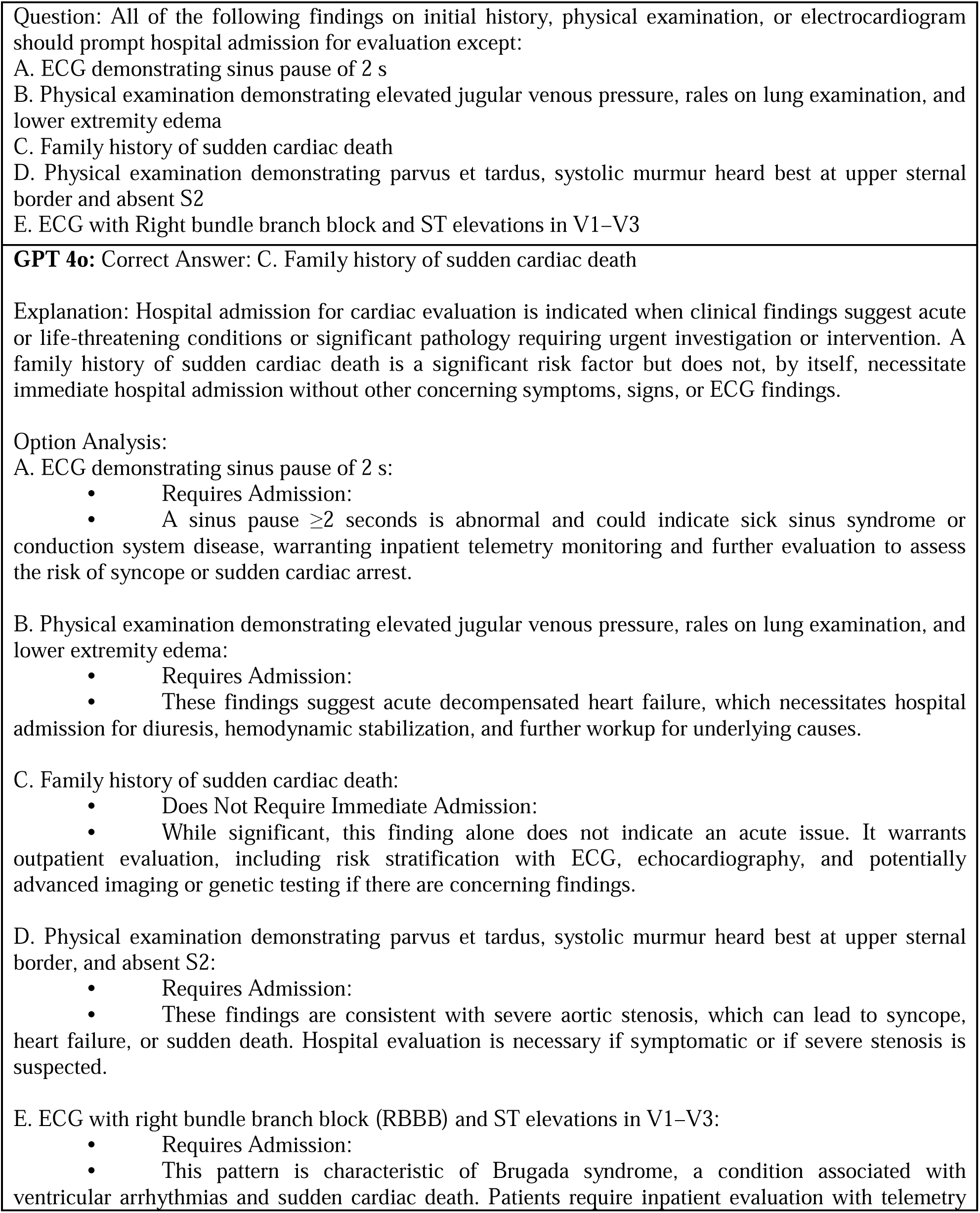
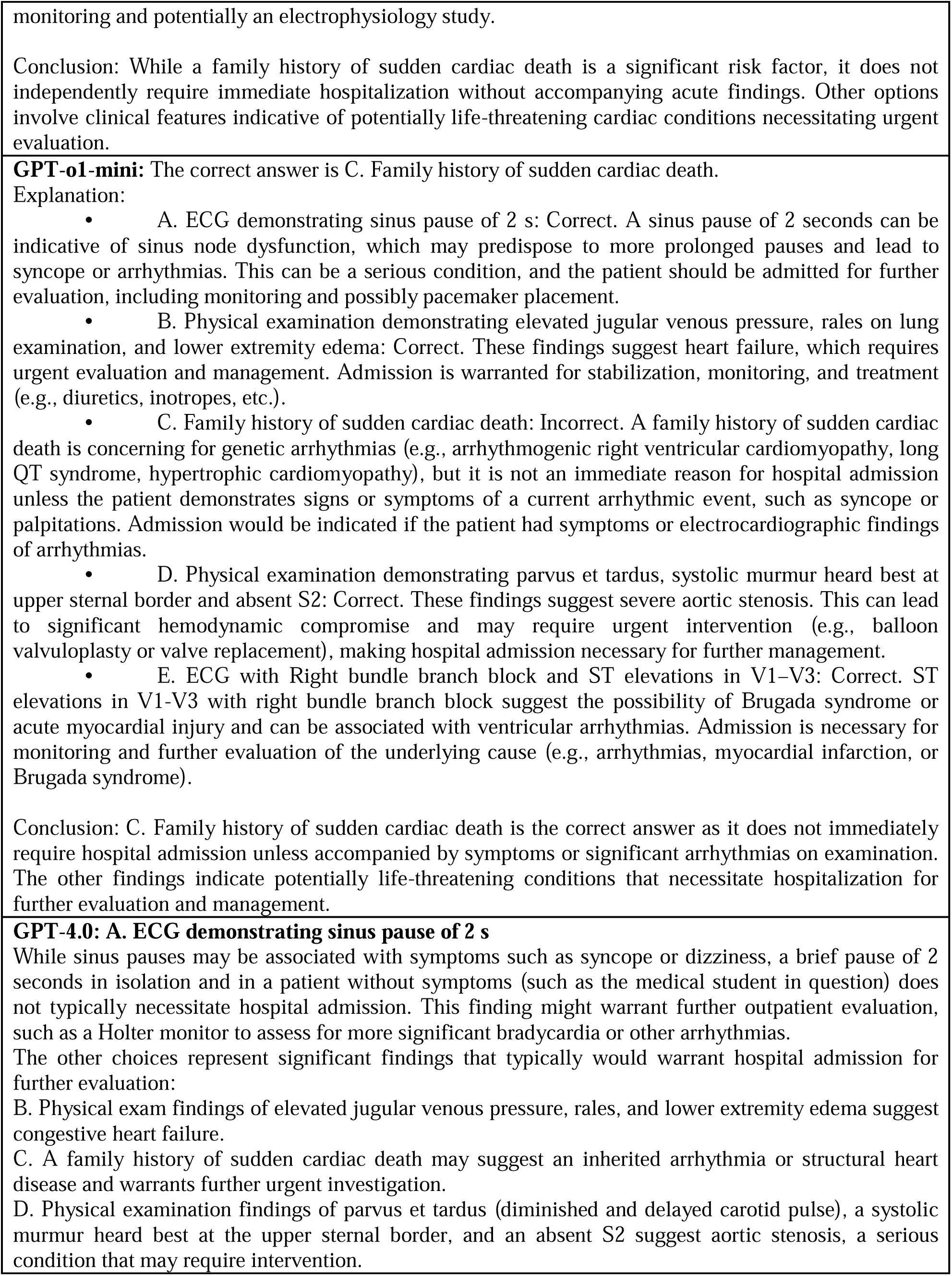
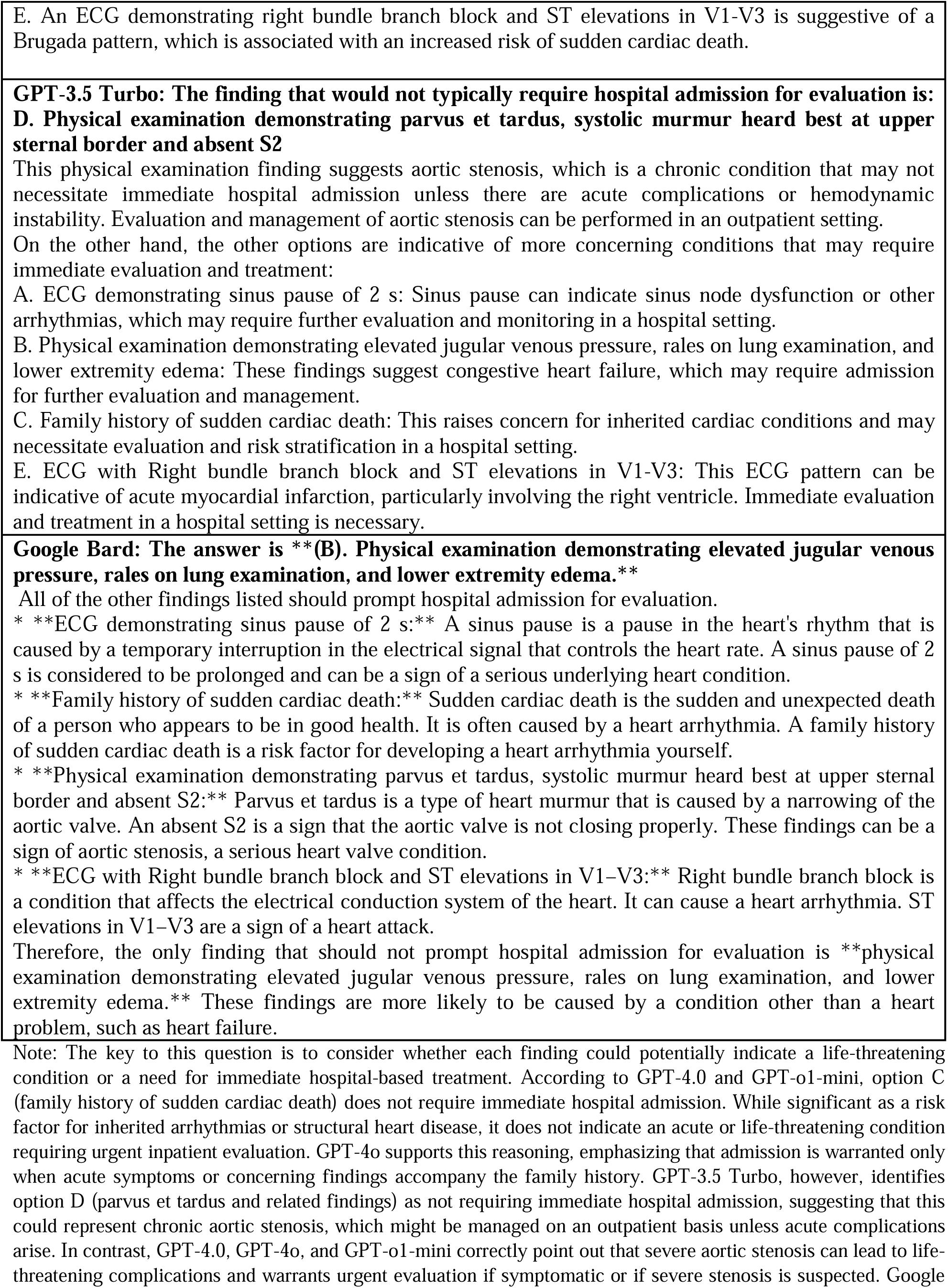
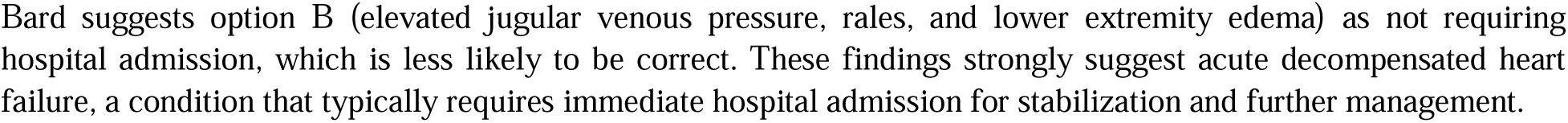
(Test set D, Question 8, Chapter 17 in. (**30**)) This table presents clinical findings (e.g. in the Emergency department) from initial history, physical examination, or electrocardiogram, and asks about the necessity for urgent hospital admission.

**Table 5.**
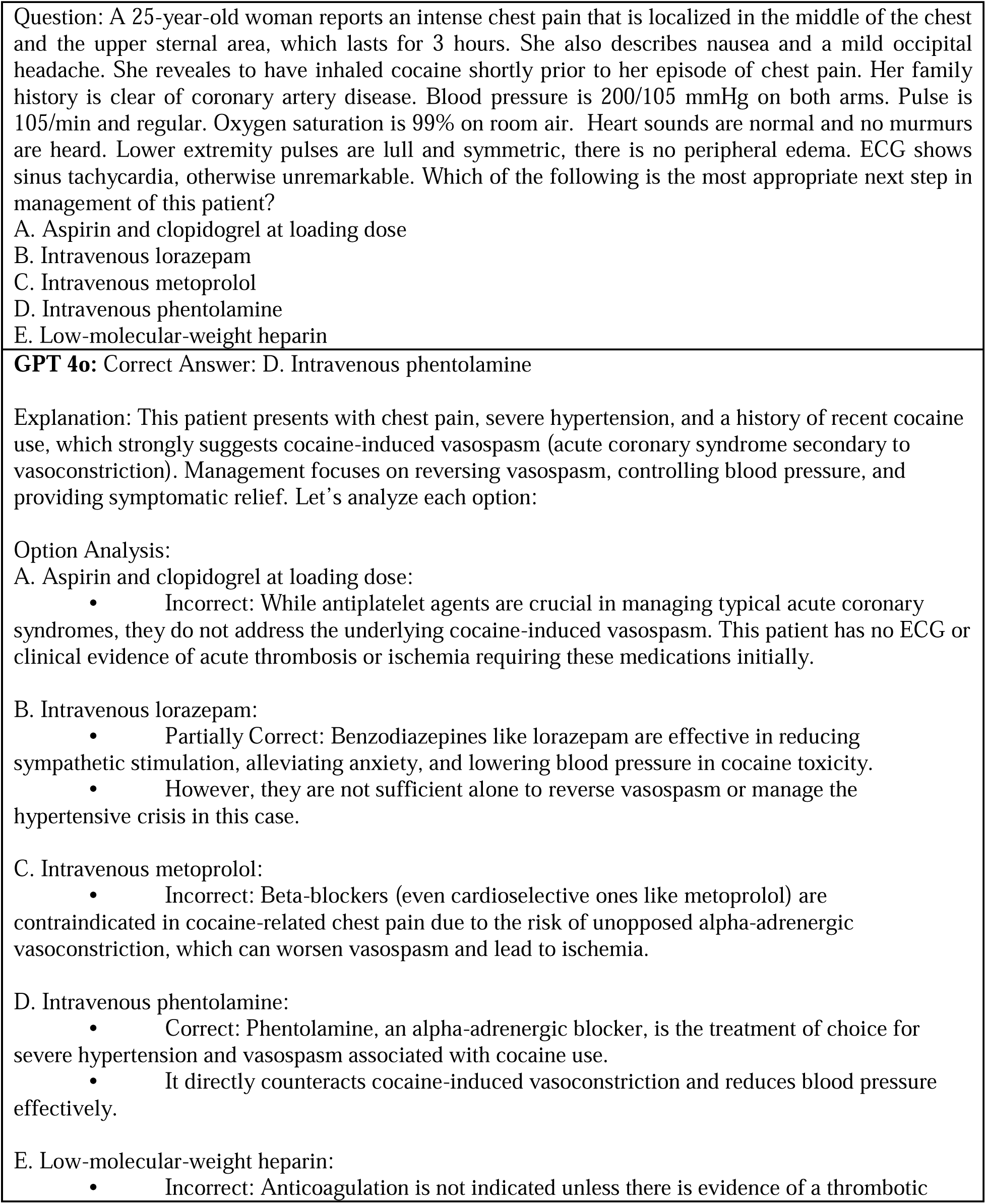
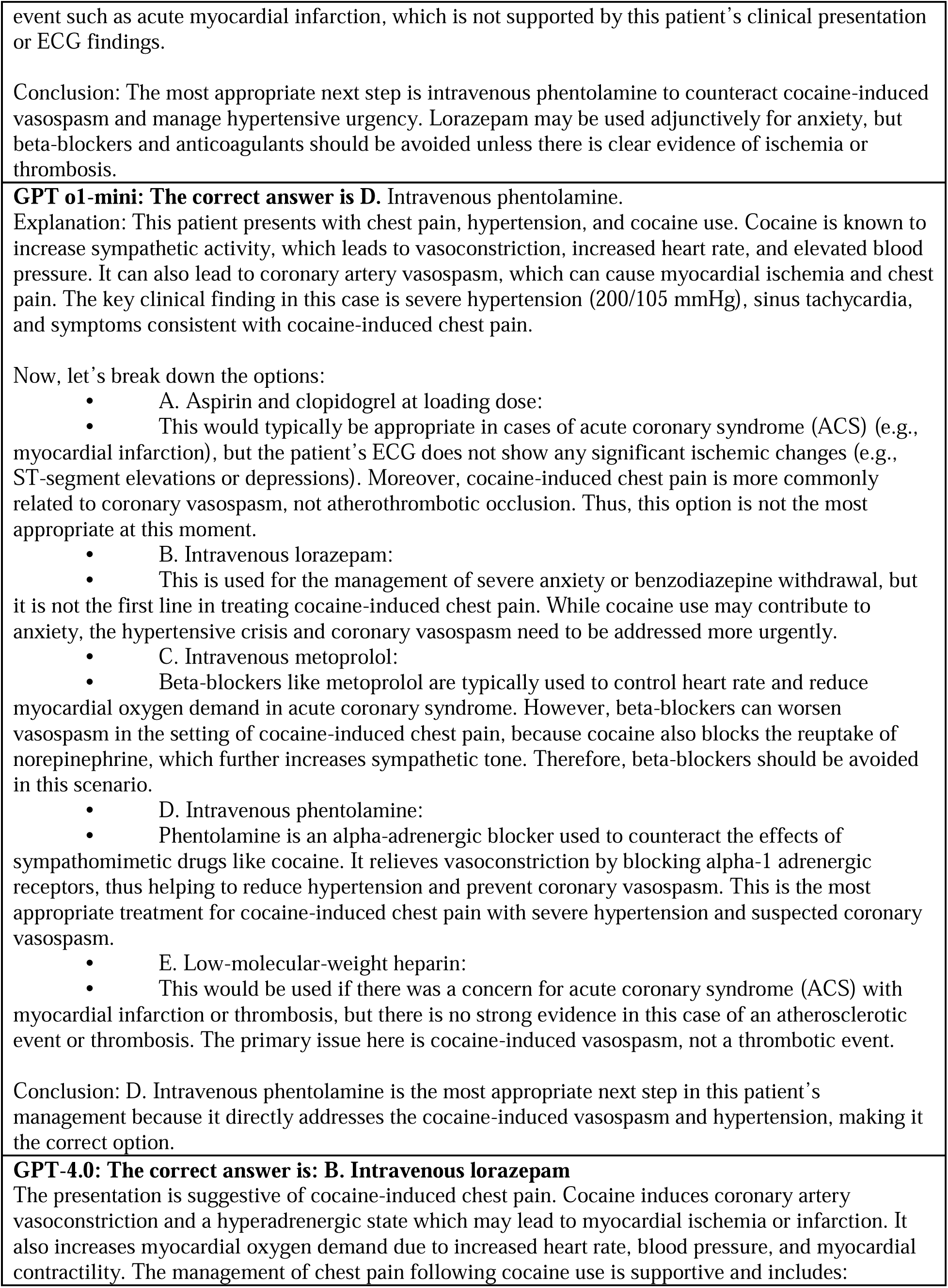
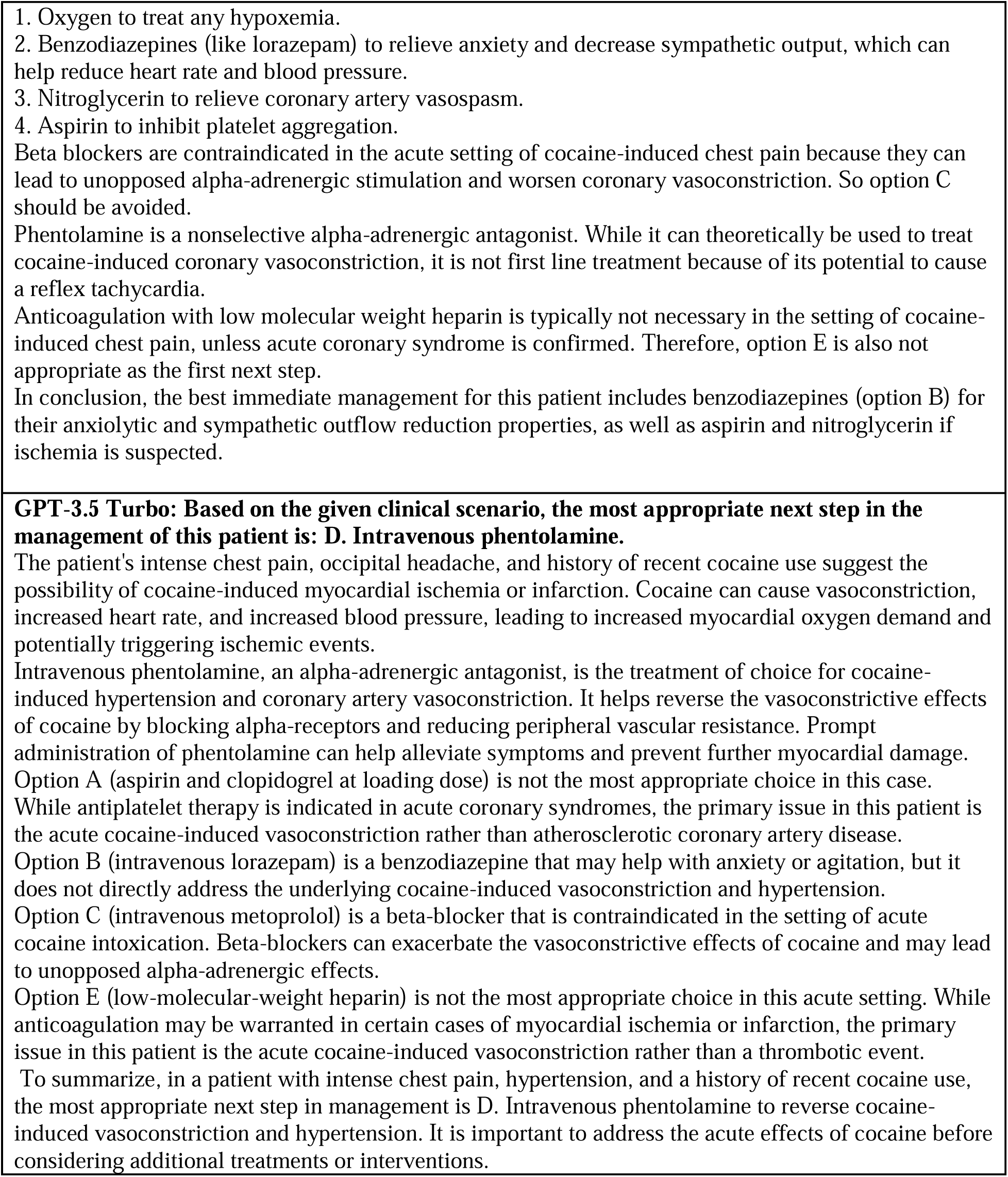
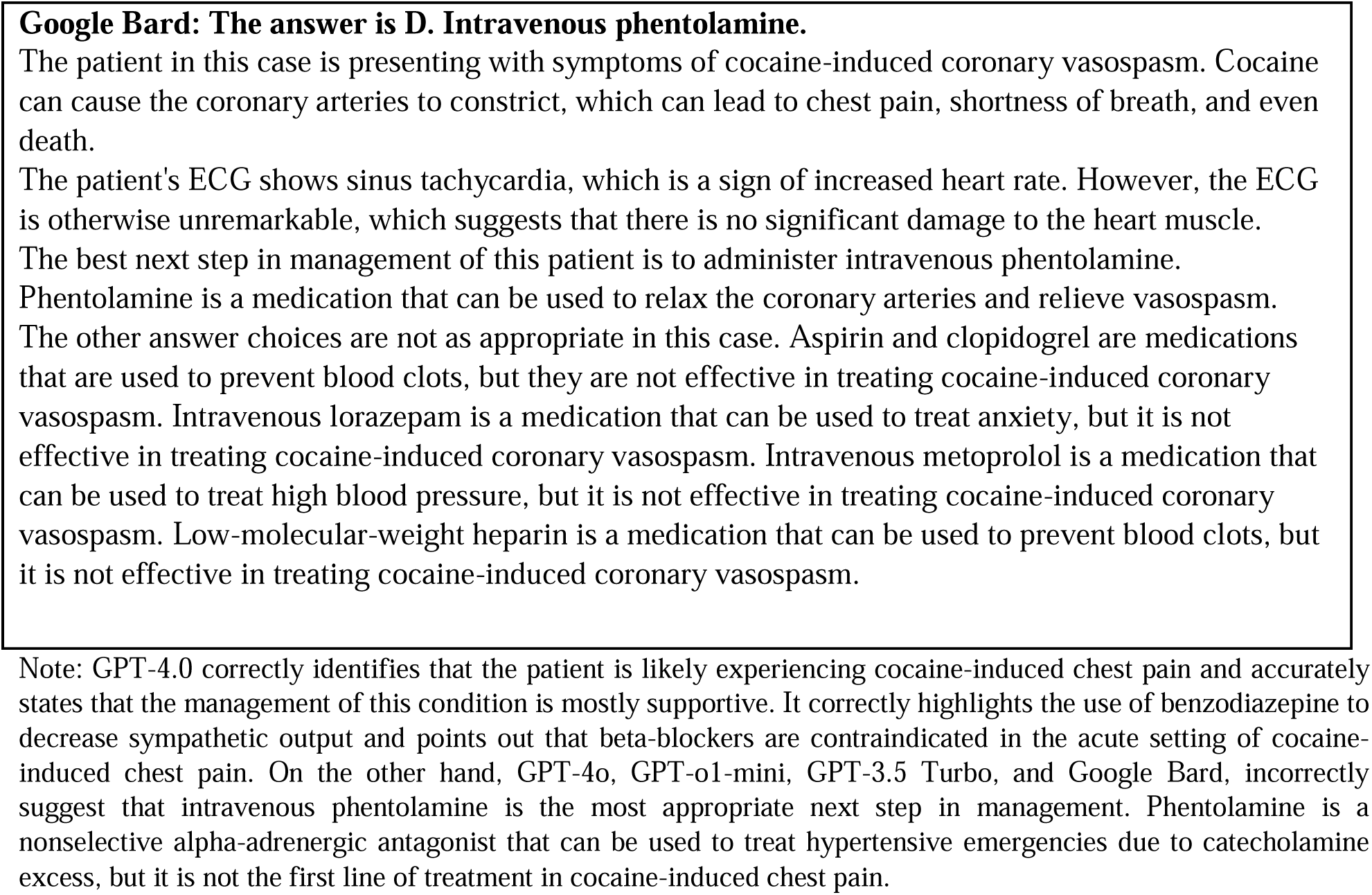
(Test set C) The table presents the case of a 25-year-old woman with chest pain, nausea, and a headache after cocaine use. It offers a detailed account of her medical condition, including vitals and examination results, with a multiple-choice question about the most suitable next step in her treatment.

**Table 6.**
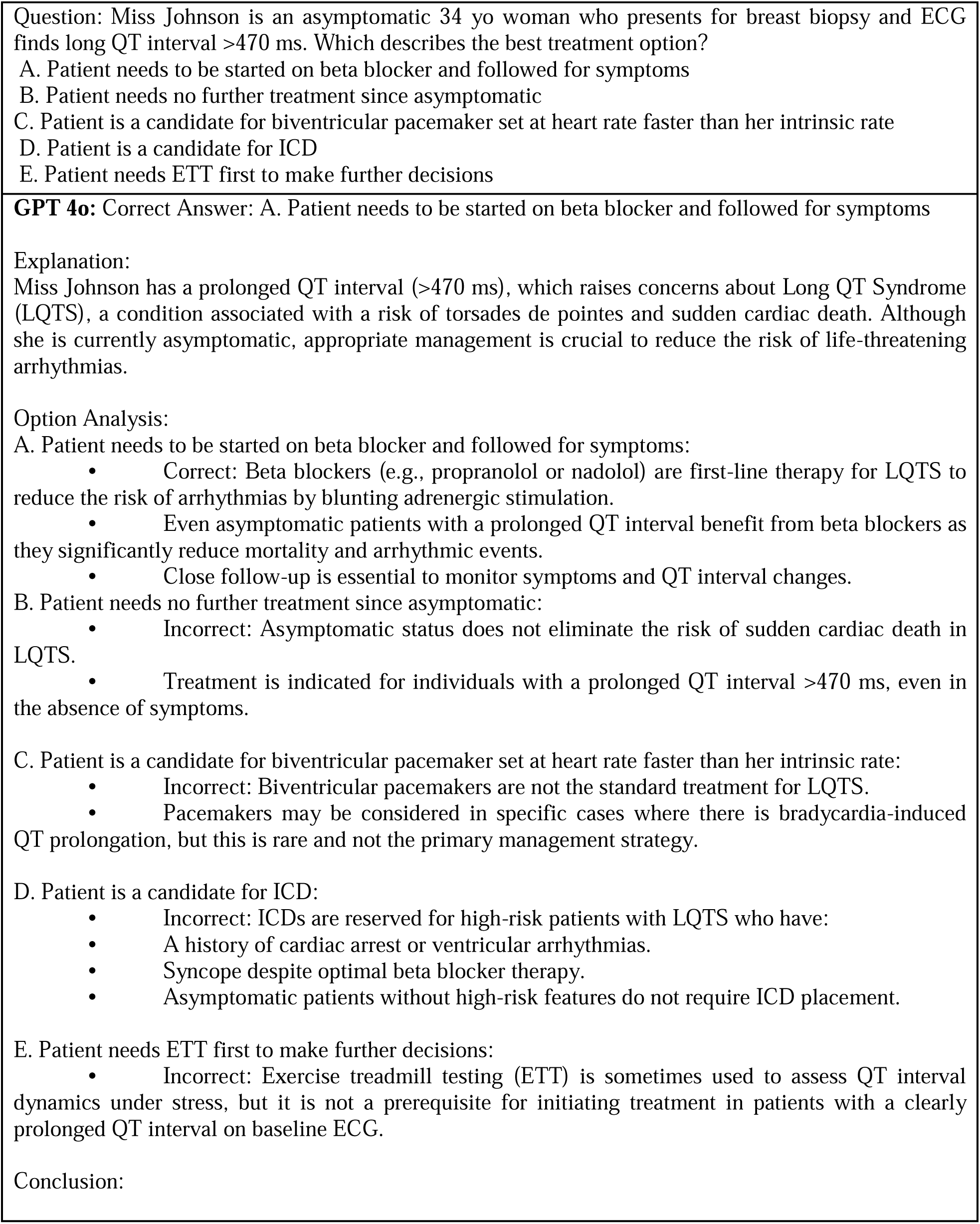
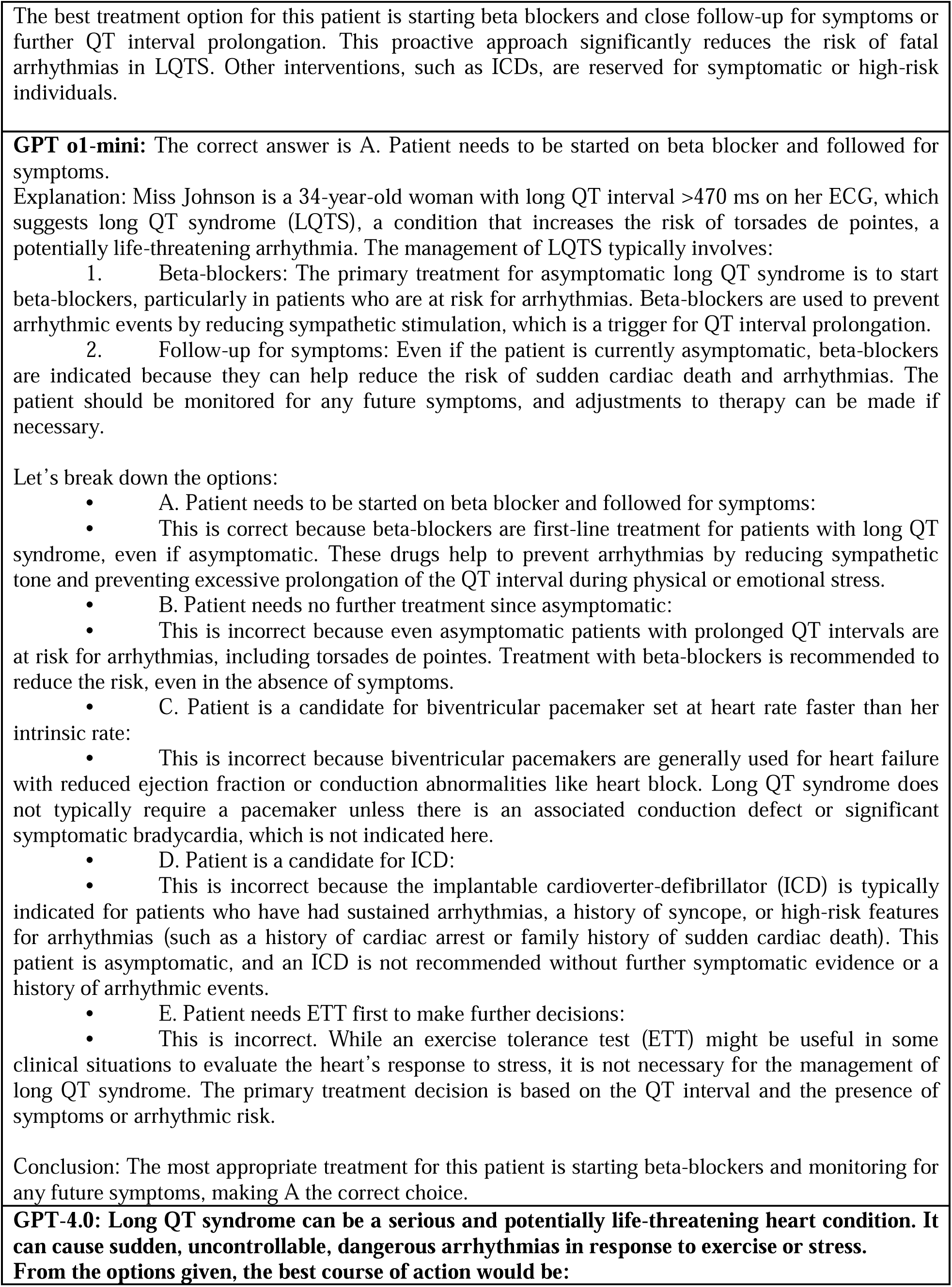
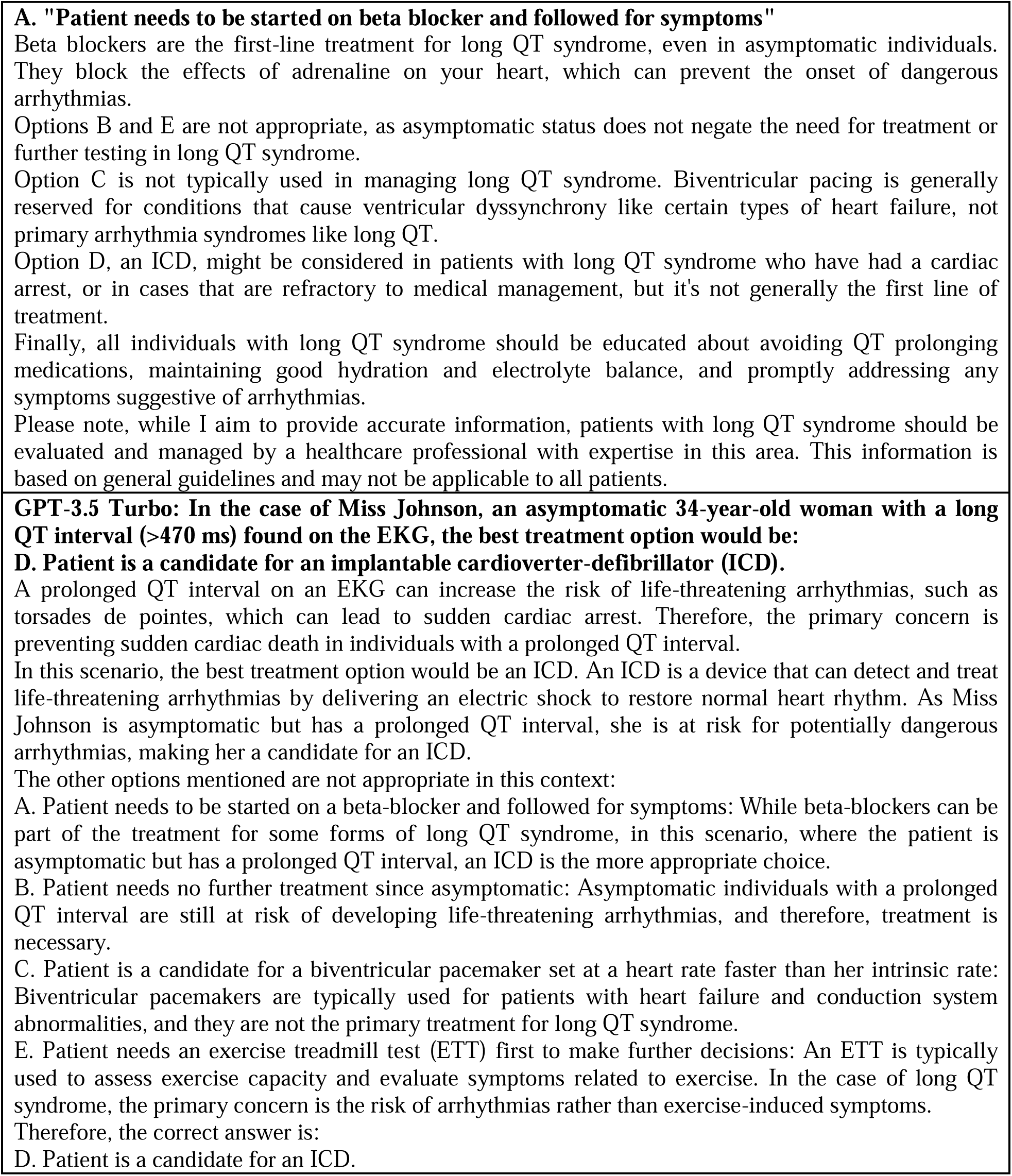
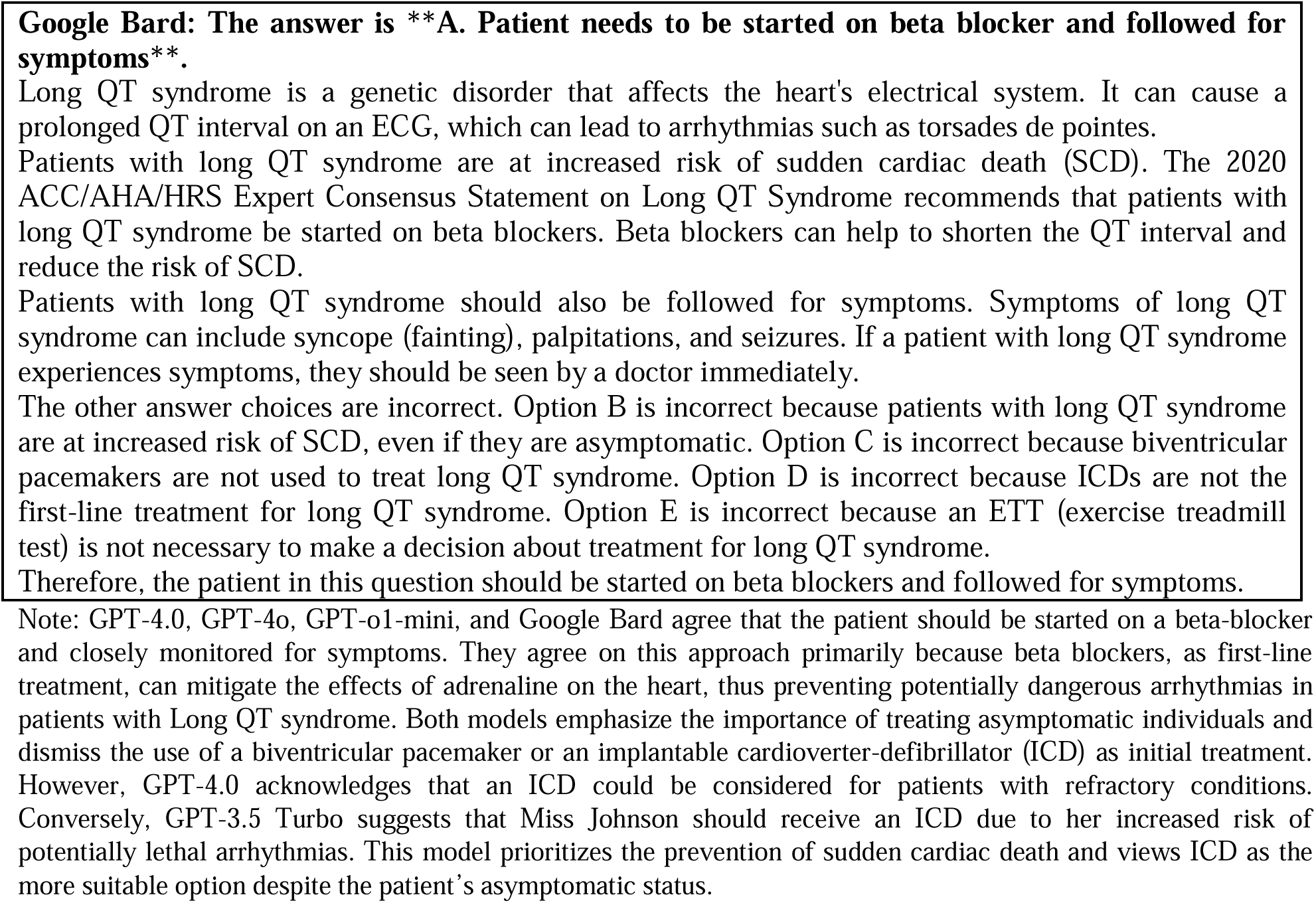
(Test set B) Provides a case of a 34-year-old asymptomatic woman with an ECG finding of a long QT interval with a multiple-choice question about the most suitable next step in her treatment.

### Statistical Analysis

A Friedman test was conducted to assess performance differences among five models—GPT-4.0, GPT-3.5 Turbo, Google Bard, GPT-4o, and GPT-o1-mini—across four test sets. For Test set A, the analysis did not reveal any statistically significant differences across the models (p = 0.112). Similarly, for Test set B, no significant differences were observed (p = 0.068). However, significant differences were found for Test set C (p = 0.004) and Test set D (p < 0.001), indicating variation in performance across models on more complex test sets.

Post-hoc pairwise comparisons provided additional insights. For Test set A, no significant differences were identified between any of the models after Bonferroni correction. On Test set B, while differences were not statistically significant, GPT-4.0 displayed a trend of higher performance compared to Google Bard and GPT-3.5 Turbo. On Test set C, GPT-4.0 significantly outperformed Google Bard (adjusted p = 0.002), but the difference between GPT-4.0 and GPT-3.5 Turbo was not statistically significant (adjusted p = 0.062). Additionally, GPT-4.0 showed no significant performance difference compared to GPT-4o or GPT-o1-mini for this test set. For Test set D, GPT-4.0 demonstrated statistically significant superiority over both Google Bard and GPT-3.5 Turbo (adjusted p < 0.001 for both). GPT-4.0 also significantly outperformed GPT-4o (adjusted p = 0.038), although no significant difference was detected between GPT-4.0 and GPT-o1-mini (adjusted p = 0.177).

## Discussion

In the following sections, we will highlight five vignettes that demonstrate specific characteristics of each LLM. We refer the reader to Appendix 1 for further selected vignettes and their interpretations. Notably, GPT-4.0, GPT-4o, GPT-o1-mini, GPT-3.5 Turbo, and Google Bard yield outputs that manifest a high level of comprehensibility, demonstrating the potential utility of these models in relaying medical information to diverse audiences.

### Model Performance and Proficiency in Medical Terminology

The models’ responses were coherent, structured, and detailed, effectively translating complex medical jargon into accessible language. GPT-4.0 consistently outperformed other models across all test sets, demonstrating a superior ability to integrate clinical information and adhere to current medical guidelines. However, the relatively smaller number of questions in test sets A and B may have reduced the statistical power, leading to a lack of significant differences for those sets despite observable performance trends. Critically, each model showed the capacity to identify and process important clinical details from the input, apply medical knowledge, and generate contextually appropriate responses.

All models included explanations for their selected answers, rationalizing why certain options were correct and others were not. For instance, as shown in Table 2, GPT-4.0 and GPT-3.5 Turbo displayed a strong grasp of cardiological terminology, accurately interpreting ECG and echocardiographic findings, and recommending appropriate procedures. Conversely, while Google Bard failed to recognize key details such as the wide QRS complex and the role of cardiac resynchronization therapy. GPT-4o and GPT-o1-mini also showed strong performance in clinical reasoning, often closely aligning with GPT-4.0, though subtle differences were evident in certain nuanced scenarios. In more challenging scenarios (Appendix 1, Set D, Question 3), GPT-4.0’s alignment with guideline-based care outshone the less consistent recommendations of Google Bard, reinforcing that complexity levels reveal pronounced model disparities.

### Variability in Clinical Relevance and Contextual Understanding

The study’s examination of clinical relevance and contextual understanding in the responses of GPT-4.0, GPT-4o, GPT-o1-mini, GPT-3.5 Turbo, and Google Bard revealed distinct variations in their ability to apply medical guidelines accurately. For instance, in the vignette addressing lipid management (Table 3), GPT-4.0, GPT-4o, and GPT-o1-mini consistently recommended adding ezetimibe to high-intensity statin therapy based on the patient’s slightly elevated LDL-C level. These models demonstrated proficiency in synthesizing the patient’s clinical data and aligning their recommendations with current medical guidelines. GPT-o1-mini, while closely matching the performance of GPT-4.0, occasionally provided slightly less detailed explanations but maintained adherence to clinical standards.

GPT-3.5 Turbo displayed a solid conceptual understanding, referencing the patient’s history, current statin therapy, and lipid profile in its explanation. However, it failed to align its final recommendation with current medical guidelines, reflecting a gap in contextual application despite clear reasoning. Conversely, Google Bard exhibited only partial contextual understanding. In the lipid management vignette, it correctly identified the patient’s acute myocardial infarction and noted that their LDL-C was above target, but it incorrectly categorized atorvastatin as a moderate-intensity statin and recommended switching to evolocumab. This suggestion is inconsistent with guideline-directed management and highlights limitations in its application of evidence-based practices.

In the vignette presented in Table 4, which involved recommending the necessity for urgent hospital admission based on clinical findings, GPT-4.0, GPT-4o, and GPT-o1-mini provided accurate and contextually relevant responses. In contrast, Google Bard not only offered an incorrect answer but also provided contradictory arguments that lacked clarity, further underscoring deficiencies in its contextual understanding of clinical scenarios. Furthermore, for cocaine-induced chest pain (Appendix 1, Table 5), GPT-4.0 and GPT-o variants selected benzodiazepines as the appropriate initial step, reflecting a prudent adherence to first-line treatments, unlike models that prematurely escalated care.

Finally, all models occasionally demonstrated a tendency toward overtreatment, at times bypassing steps in guideline-oriented procedures. This inclination often involved proposing more advanced diagnostic or therapeutic approaches that were unnecessary or anticipatory. As shown in Table 5, GPT-4.0, GPT-4o, and GPT-o1-mini avoided this pitfall by correctly recommending intravenous benzodiazepines as the first-line treatment for cocaine-induced chest pain. This approach aligns with clinical guidelines that emphasize the role of benzodiazepines in reducing sympathetic stimulation, which is the underlying mechanism in such cases. In contrast, GPT-3.5 Turbo and Google Bard opted for immediate treatment with phentolamine, which, while effective for addressing hypertensive emergencies in cocaine toxicity, is not the first-line recommendation for chest pain management in this context.

### Reliability and Response Consistency Across Models

Among the five models analyzed, GPT-4.0, GPT-4o, and GPT-o1-mini demonstrated the highest levels of reliability, consistently applying clinical guidelines and generating responses aligned with evidence-based practices. GPT-3.5 Turbo and Google Bard, on the other hand, exhibited notable fluctuations in their answers when presented with the same query at different times. Notably, when GPT-3.5 Turbo and Google Bard produced an incorrect response initially, their revised answers upon subsequent queries often remained incorrect. Additionally, GPT-3.5 Turbo displayed a unique tendency to endorse one response initially and shift to an alternate response by the conclusion of its output, creating confusion about its position. Google Bard also exhibited instances of internal contradiction, where it simultaneously justified and dismissed certain options within the same response (as observed in Table 4). Conversely, the GPT-4o series models showed a marked improvement in consistency, with minimal fluctuations or contradictions in their reasoning, further highlighting their reliability compared to GPT-3.5 Turbo and Google Bard.

### Validation with Clinical Trials and Guidelines

During our evaluation, GPT-4.0, GPT-4o, and GPT-o1-mini consistently applied established guidelines—such as AHA/ACC recommendations for lipid management (e.g., adding ezetimibe for elevated LDL-C in Table 3) and expert consensus on arrhythmia care (e.g., beta-blockers for long QT in Table 6)—with a high degree of accuracy. However, it should be noted that the responses from GPT-4.0 and related models are predicated on medical literature and guidelines available until their respective knowledge cutoffs in 2023, potentially limiting their applicability to very recent advances. Google Bard (e.g. Table 6), while occasionally referencing an Expert Consensus Statement, applied these guidelines less consistently. In contrast, GPT-3.5 Turbo deviated from the guidelines by recommending an implantable cardioverter-defibrillator (ICD) without considering beta-blockers as the first-line treatment, showcasing a gap in guideline alignment.

Lastly, none of the models demonstrated the ability to incorporate or reference ongoing clinical trials within their responses. This limitation highlights an opportunity for future models to integrate real-time clinical trial data for enhanced decision-making and relevance.

### Speed and Efficiency

Assessing the speed and efficiency of the models is inherently challenging due to variability in execution time influenced by user load and server conditions. GPT-4.0, GPT-4o, GPT-o1-mini, and GPT-3.5 Turbo exhibited steady response generation, with incremental streaming of outputs, which helped reduce perceived waiting time and allowed for real-time assessment of answers. In contrast, Google Bard holds back its outputs, delivering the complete text following a slight delay.

### Adaptability to Different Sub-Domains

The robust performance of GPT-4.0, GPT-4o, and GPT-o1-mini, alongside the solid contributions of GPT-3.5 Turbo and Google Bard in various cardiological scenarios, highlights their potential applicability to other medical specialties. These models demonstrate a capacity to synthesize and present complex medical information concisely and accurately, qualities that are invaluable across diverse medical sub-domains.LLMs can quickly analyze large sets of data, including electronic health records and genomic data (8). This allows them to identify patterns not apparent to humans, recognize potential risk factors, and even provide outcome predictions (9). However, one must also consider the breadth and depth of the model’s training data. As their capabilities are inherently shaped by the corpus they were trained on, their efficacy across different medical domains may vary based on the representation of those domains in the original dataset. In a hospital environment, they have the potential to serve as a medium for repetitive tasks, such as assisting with writing discharge letters by summarizing a patient’s hospital stay after reading their medical records or providing further recommendations (3,7). A study from 2020 demonstrated that ML, integrating clinical parameters with coronary artery calcium and automated epicardial adipose tissue quantification, significantly improved the prediction of myocardial infarction and cardiac death compared with standard clinical risk assessment (23). Therefore, while we may anticipate a degree of adaptability, evaluating their performance individually within each target sub-domain is advisable to ensure precision and validity (32–35).

On the other hand, although these models demonstrate proficiency in generating linguistically precise content, they do not have the capability to comprehend or internalize knowledge of the world in a manner akin to human cognition. Clinical decision-making requires the synthesis of evidence-based medicine, guidelines, and sound clinical judgment (10). Intuition, based on knowledge and care experience, also plays a crucial role (11,12). The rapid evolution of LLMs and the burgeoning variety of available models warrant systematic comparative analyses or benchmarking (13–16). Consensus indicates that LLMs possess the capability to aid in clinical case resolution; however, their application necessitates caution due to inherent imprecision and a propensity for disseminating misinformation (17,18). Overseeing the impending incorporation of this AI into daily medical practice is essential to ensure the effective management of knowledge sources and their application.

### Potential for Personalization

Personalization is a crucial attribute for LLMs, focusing on integrating and interpreting user-specific information into their responses (34,35). In this study, we provide ‘Dubravka’, a custom-made mobile application, that interfaces with the GPT-3.5 Turbo API (Application Programming Interface). This application’s primary function is to incorporate user-specific medical data to tailor the LLM’s responses to individual health profiles. To achieve this, users are prompted to input basic medical information during their initial interaction with the application. This data is then stored locally on the phone and included as a hidden parameter in the conversation, instructing the LLM to moderate its responses specifically to each patient’s medical history, drug therapy, and lifestyle elements. This setup allows the LLM to process individual health data and generate advice based on the user’s prompt and the provided medical information, with a primary emphasis on diet, cardiovascular prevention, and lifestyle recommendations. We release this simple prototype in the hope that it will serve as a potential use case for the development of more sophisticated professional-level applications. Additionally, the complete source code is publicly available for those interested in further development or utilization of this framework. Notably, the open-source architecture allows users to modify the application to interface with more advanced models, such as GPT-4, enabling customization based on individual preferences and budgetary constraints. In Figure 2, we present the user interface of ‘Dubravka’ as it appears to a patient with recent deep vein thrombosis, along with her question about the consistent use of compressive stockings. In Appendix 2, we highlight 10 typical patient profiles and their inquiries about their medical conditions. The answers were obtained with the temperature parameter set to zero. Those synthetic patients have been proposed by the authors of this research, reflecting the typical patients from our practice in a high-volume tertiary center. Exploring the Dubravka prototype demonstrates that LLMs have the potential to be integrated into patient-facing tools, streamlining patient education, medical history summaries, and prevention guidance. Although currently focused on lifestyle and prevention, this approach highlights LLMs’ adaptability for personalized clinical support.

**Figure 2.**
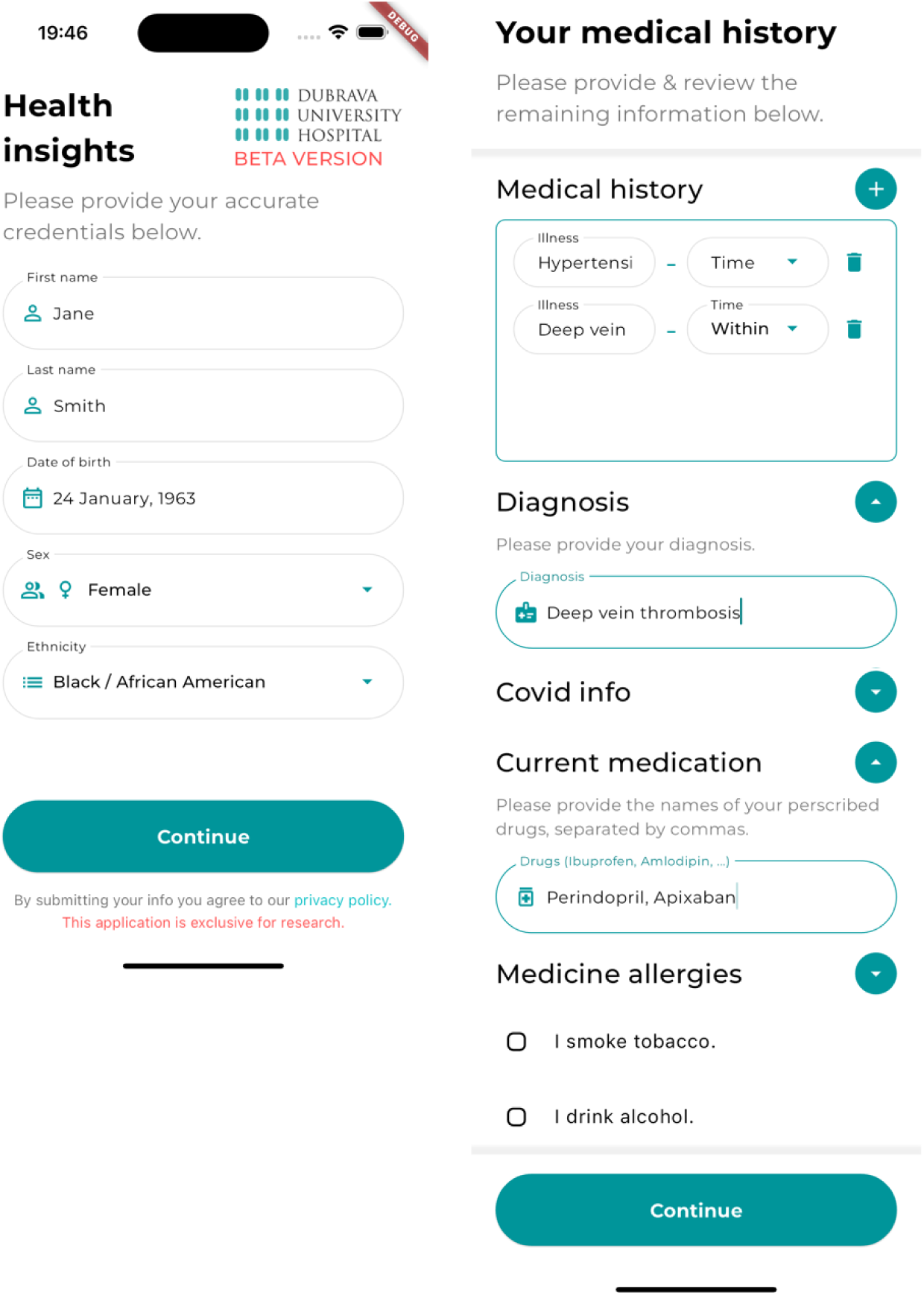
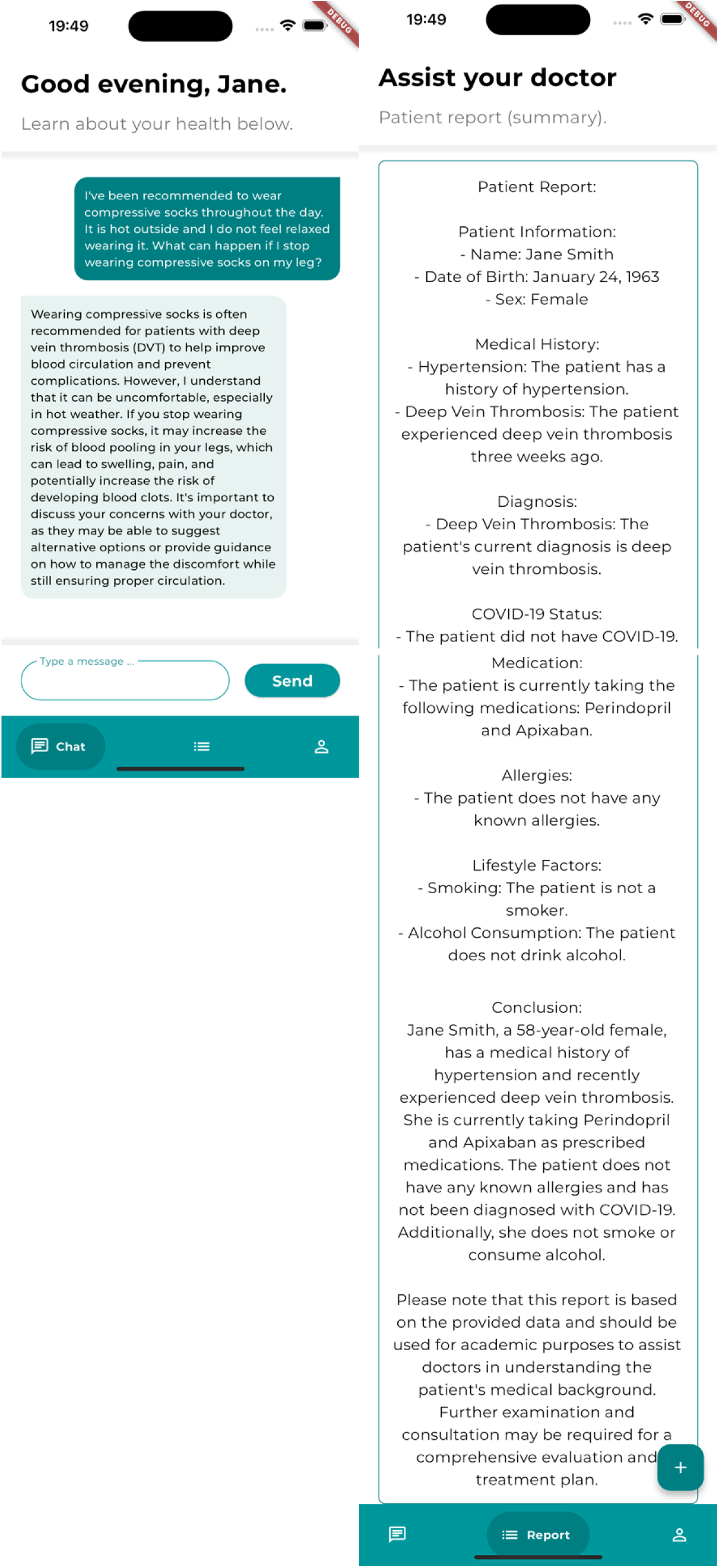
The interface of the ‘Dubravka’ mobile application. Images represent the user interface of the ‘Dubravka’ mobile application, specifically designed within this research. This digital health tool incorporates the GPT-3.5 Turbo Application Programming Interface (API), thus enabling the LLM to interact with unique health data from individual users. Consequently, the application has the potential to generate tailored advice, with an emphasis on areas such as dietary habits, lifestyle modifications, and cardiovascular prevention strategies. In addition, the application includes a feature that facilitates the summarization of a patient’s medical history, potentially aiding in the communication between patients and healthcare professionals. The images in the first row depict the information that the patient enters, while the second row shows communication with the LLM, and the essential medical history summarized by the LLM based on the given data (first row). The images have been cropped to contain only the relevant information.

## Limitations

This study, while comprehensive in its assessment of three distinct LLMs, through both horizontal and vertical evaluations, carries several limitations. Firstly, its cross-sectional (observational) design, employing a fixed set of questions with predetermined correct answers, may not fully capture the dynamic and multifaceted nature of clinical decision-making in real-world cardiology. Moreover, while the questions contained information derived from investigative methods such as ECGs and sonography, LLMs were not asked to interpret images directly. Secondly, it is worth noting that the power of a statistical test, its capacity to detect a genuine effect when it exists, can be influenced by the sample size. This is particularly true for the initial two test sets, where the fewer number of questions might have reduced the sensitivity in detecting significant differences in model performance.

Additionally, the lack of clear and concise guidelines for setting the ‘temperature’ of LLMs is a notable limitation. The study’s methodology, which relied on the knowledge of a clinical cardiologist rather than an IT expert, reflects a real-world scenario where clinicians are typically not versed in adjusting the LLMs’ parameter settings, potentially leading to variations in user experience. However, current evidence supports the notion that temperature variations reflect the diversity in phrasing of the answer, rather than the representation of medical knowledge encoded in the model (29).

Another critical aspect concerns the financial accessibility of some LLMs. Our study relied on GPT-4, GPT-4o and GPT-o1-mini a paid models, suggesting that routine clinical use might be limited to medical staff who can afford it. This raises questions about equitable access to such advanced AI tools in diverse clinical settings.

The findings of this study are also contingent on the models’ knowledge as of their last training update (as of September 2021). Considering the rapid evolution in AI capabilities and medical knowledge, the relevance and accuracy of these findings are subject to change over time.

Lastly, the study did not address the ethical implications and practical challenges of integrating AI in clinical settings, including concerns about patient privacy, data security, and the impact on the physician-patient relationship. While the models demonstrated adaptability, their performance in other medical fields may vary depending on the representation of those domains in their training datasets.

## Conclusion

This study benchmarks the performance of five large language models (LLMs)—GPT-4.0, GPT-4o, GPT-o1-mini, GPT-3.5 Turbo, and Google Bard—on their ability to respond to diverse clinical scenarios in cardiology. The evaluation focused on understanding medical terminology, clinical relevance, contextual accuracy, and guideline adherence.

Among these models, GPT-4.0 consistently demonstrated superior performance, showing high reliability, accuracy, and compliance with contemporary medical guidelines, making it the most suitable for real-world clinical applications. GPT-4o and GPT-o1-mini also performed well, closely following GPT-4.0, while GPT-3.5 Turbo displayed a solid grasp of medical concepts but occasionally diverged from guideline-based recommendations. Google Bard, despite some strengths, showed inconsistencies in contextual understanding and a tendency to deviate from established guidelines.

This study underscores the importance of continual validation of these models across medical specialties, ensuring they remain updated with the latest clinical guidelines and evidence. Moreover, the incorporation of open-source frameworks offers a pathway for cost-effective solutions, enabling broader accessibility while allowing customization for specific clinical settings.

As AI continues to transform healthcare, this research reinforces the critical role of LLMs as tools to augment, not replace, medical professionals. By approaching these technologies with critical engagement and a commitment to ongoing refinement, we can harness their potential to enhance patient care and support clinical decision-making.

## Data Availability

All data produced in the present study are available upon reasonable request to the authors

## Acknowledgments

We would also like to extend our gratitude to Antonio Butigan for his contribution to this study. His technical expertise and commitment to excellence were pivotal in the programming of the ‘Dubravka’ mobile application. We gratefully acknowledge the Luxembourg School of Business for providing the technological support for this research. This work was partially supported by the Croatian Science Foundation under project numbers HRZZ-MOBODL-2023-08-7617 and IP-2022-10-7261 (ADESO), and by the University of Zagreb Support, Grant number 10106-24-1505. Additionally, the author(s) would like to acknowledge the contribution of the COST Action CA21169, supported by COST (European Cooperation in Science and Technology).

## Footnotes

1 Parameter settings and recommendations could be found on: https://platform.openai.com/docs/api-reference/chat/create

2 https://jupyter.org/

## Notes

### Competing Interest Statement

The authors have declared no competing interest.

### Funding Statement

This study did not receive any funding.

### Summary of Updates

Figure 1 revised and updated with GPT o-series.

